# The statistical analysis of daily data associated with different parameters of the New Coronavirus COVID-19 pandemic in Georgia and their monthly interval prediction from September 1, 2021 to December 31, 2021

**DOI:** 10.1101/2022.01.16.22269373

**Authors:** Avtandil G. Amiranashvili, Ketevan R. Khazaradze, Nino D. Japaridze

## Abstract

The lockdown introduced in Georgia on November 28, 2020 contributed to positive trends in the spread of COVID-19 until February - the first half of March 2021. Then, in April-May 2021, the epidemiological situation worsened significantly, and from June to the end of December COVID - situation in Georgia was very difficult.

In this work results of the next statistical analysis of the daily data associated with New Coronavirus COVID-19 infection of confirmed (C), recovered (R), deaths (D) and infection rate (I) cases of the population of Georgia in the period from September 01, 2021 to December 31, 2021 are presented. It also presents the results of the analysis of monthly forecasting of the values of C, D and I. As earlier, the information was regularly sent to the National Center for Disease Control & Public Health of Georgia and posted on the Facebook page https://www.facebook.com/Avtandil1948/.

The analysis of data is carried out with the use of the standard statistical analysis methods of random events and methods of mathematical statistics for the non-accidental time-series of observations. In particular, the following results were obtained.

Georgia’s ranking in the world for Covid-19 monthly mean values of infection and deaths cases in investigation period (per 1 million population) was determined. Among 157 countries with population ≥ 1 million inhabitants in October 2021 Georgia was in the 4 place on new infection cases, and in September - in the 1 place on death. Georgia took the best place in terms of confirmed cases of diseases (thirteenth) in December, and in mortality (fifth) - in October.

A comparison between the daily mortality from Covid-19 in Georgia from September 01, 2021 to December 31, 2021with the average daily mortality rate in 2015-2019 shows, that the largest share value of D from mean death in 2015-2019 was 76.8 % (September 03, 2021), the smallest 18.7 % (November 10, 2021).

As in previous work [9,10] the statistical analysis of the daily and decade data associated with coronavirus COVID-19 pandemic of confirmed, recovered, deaths cases and infection rate of the population of Georgia are carried out. Maximum daily values of investigation parameters are following: C = 6024 (November 3, 2021), R = 6017 (November 15, 2021), D = 86 (September 3, 2021), I = 12.04 % (November 24, 2021). Maximum mean decade values of investigation parameters are following: C = 4757 (1 Decade of November 2021), R = 4427 (3 Decade of November 2021), D = 76 (2 Decade of November 2021), I = 10.55% (1 Decade of November 2021).

It was found that as in spring and summer 2021 [9,10], from September to December 2021 the regression equations for the time variability of the daily values of C, R, D and I have the form of a tenth order polynomial.

Mean values of speed of change of confirmed -V(C), recovered - V(R), deaths - V(D) and infection rate V(I) coronavirus-related cases in different decades of months for the indicated period of time were determined. Maximum mean decade values of investigation parameters are following: V(C) = +139 cases/day (1 Decade of October 2021), V(R) = +124 cases/day (3 Decade of October 2021), V(D) = +1.7 cases/day (3 Decade of October 2021), V(I) = + 0.20 %/ day (1 decades of October 2021).

Cross-correlations analysis between confirmed COVID-19 cases with recovered and deaths cases shows, that from September 1, 2021 to November 30, 2021 the maximum effect of recovery is observed on 12 and 14 days after infection (CR=0.77 and 0.78 respectively), and deaths - after 7, 9, 11, 13 and 14 days (0.70≤CR≤0.72); from October 1, 2021 to December 31, 2021 - the maximum effect of recovery is observed on 14 days after infection (RC=0.71), and deaths - after 9 days (CR=0.43). In Georgia from September 1, 2021 to November 30, 2021 the duration of the impact of the delta variant of the coronavirus on people (recovery, mortality) could be up to 28 and 35 days respectively; from October 1, 2021 to December 31, 2021 - up to 21 and 29 days respectively.

Comparison of daily real and calculated monthly predictions data of C, D and I in Georgia are carried out. It was found that in investigation period of time daily and mean monthly real values of C, D and I practically fall into the 67% - 99.99% confidence interval of these predicted values.

Traditionally, the comparison of data about C and D in Georgia (GEO) with similar data in Armenia (ARM), Azerbaijan (AZE), Russia (RUS), Turkey (TUR) and in the World (WRL) is also carried out.

## 1. Introduction

Two years have passed since the outbreak of the new coronavirus (COVID-19) in China, which was recognized on March 11, 2020 as a pandemic due to its rapid spread in the World [1]. During this period of time, despite the measures taken (including vaccination), several strains of this virus have appeared. The overall level of morbidity and mortality in many countries of the world is still quite high. Scientists and specialists of various disciplines from all over the world continue intensive research of this unprecedented phenomenon (including in Georgia [2-11]), rendering their all possible assistance to epidemiologists.

In particular, in our works [6-11], it was noted that specialists in the field of physical and mathematical sciences make an important contribution to research on the spread of the new coronavirus COVID-19. Works on statistical analysis [2-5, 12-15], forecasting [16-32], forecasting systematization [33,34], spatial-temporary modeling of the spread of the new coronavirus [35-38] etc. was actively continuing in 2021.

This work is a continuation of the researches [7-11].

In this work results of a statistical analysis of the daily data associated with New Coronavirus COVID-19 infection of confirmed (C), recovered (R), deaths (D) and infection rate (I) cases of the population of Georgia in the period from September 01, 2021 to December 31, 2021 are presented. It also presents the results of the analysis of monthly forecasting of the values of C, D and I. The information was regularly sent to the National Center for Disease Control & Public Health of Georgia and posted on the Facebook page https://www.facebook.com/Avtandil1948/.

The comparison of data about C and D in Georgia with similar data in Armenia, Azerbaijan, Russia, Turkey and in the world is also carried out.

We used standard methods of statistical analysis of random events and methods of mathematical statistics for non-random time series of observations [7-11, 39-41].

## 2. Study areas, material and methods

The study area: Georgia. Data of John Hopkins COVID-19 Time Series Historical Data (with US State and County data) [https://www.soothsawyer.com/john-hopkins-time-series-data-with-us-state-and-county-city-detail-historical/; https://data.humdata.org/dataset/total-covid-19-tests-performed-by-country] and https://stopcov.ge about daily values of confirmed, recovered, deaths and infection rate coronavirus-related cases, from September 01, 2021 to December 31, 2021 are used. The work also used data of National Statistics Office of Georgia (Geostat) on the average monthly total mortality in Georgia in January-December 2015-2019 [https://www.geostat.ge/en/]. In the proposed work the analysis of data is carried out with the use of the standard statistical analysis methods of random events and methods of mathematical statistics for the non-accidental time-series of observations [7-11, 39-41].

The following designations will be used below: Mean – average values; Min – minimal values; Max - maximal values; Range – Max-Min; St Dev - standard deviation; σ_m_ - standard error; C_V_ = 100·St Dev/Mean – coefficient of variation, %; R^2^ – coefficient of determination; r – coefficient of linear correlation; CR – coefficient of cross correlation; Lag = 1, 2…60 Day; KDW – Durbin-Watson statistic; Calc – calculated data; Real - measured data; Dec1, Dec2, Dec3 – numbers of the month decades; α - the level of significance; C, R, D - daily values of confirmed, recovered and deaths coronavirus-related cases; V(C), V(R) and V(D) - daily values of speed of change of confirmed, recovered and deaths coronavirus-related cases (cases/day); I - daily values of infection rate (or positive rate) coronavirus-related cases (100· C/number of coronavirus tests performed); V(I) - daily values of speed of change of I; DC – deaths coefficient, % = (100· D/C). Official data on number of coronavirus tests performed are published from December 05, 2020 [https://stopcov.ge].

The statistical programs Data Fit 7, Mesosaur and Excel 16 were used for calculations.

The curve of trend is equation of the regression of the connection of the investigated parameter with the time at the significant value of the determination coefficient and such values of KDW, where the residual values are accidental. If the residual values are not accidental the connection of the investigated parameter with the time we will consider as simply regression.

The calculation of the interval prognostic values of C, D and I taking into account the periodicity in the time-series of observations was carried out using Excel 16 (the calculate methodology was description in [7]). The duration of time series of observations to calculating monthly forecasts of C and I values was as follows: for September - 120 days, for October - 122 days, for November - 118 days, for December - 127 days, periodicity - 7 days. The duration of time series of observations to calculating monthly forecasts of D values was as follows: for September - 119 days, periodicity - 8 days; for October - 124 days, periodicity - 22 days; for November - 118 days, periodicity - 8 days; for December - 127 days, periodicity - 14 days.

67%…99.99%_Low - 67% 99.99% lower level of confidence interval of prediction values of C, D and I; 67%…99.99%_Upp - 67% 99.99% upper level of confidence interval of prediction values of C, D and I.

In the Table 1 [8] the scale of comparing real data with the predicted ones and assessing the stability of the time series of observations in the forecast period in relation to the pre-predicted one (period for prediction calculating) is presented.

**Table 1.** The statistical characteristics of Covid-19 confirmed cases to 1 million populations in Georgia, neighboring countries (Armenia, Azerbaijan, Russia, Turkey) and World from September 1, 2021 to December 31, 2021 (r_min_ = ± 0.18, α = 0.05).

| Parameter | GEO | ARM | AZE | RUS | TUR | WRL |
| --- | --- | --- | --- | --- | --- | --- |
| Mean | 843 | 284 | 154 | 197 | 298 | 73 |
| Min | 178 | 20 | 0 | 117 | 0 | 39 |
| Max | 1616 | 877 | 716 | 276 | 898 | 248 |
| Range | 1438 | 857 | 716 | 159 | 898 | 208 |
| Median | 806 | 226 | 137 | 198 | 296 | 65 |
| St Dev | 358 | 214 | 119 | 49 | 84 | 33 |
| $\sigma_m$ | 32.6 | 19.4 | 10.8 | 4.4 | 7.6 | 3.0 |
| $C_v$ (%) | 42.5 | 75.2 | 77.1 | 24.8 | 28.1 | 45.7 |
| Correlation Matrix |  |  |  |  |  |  |
| GEO | 1 | **0.50** | **0.23** | **0.67** | 0.01 | -0.14 |
| ARM | **0.50** | 1 | **0.23** | **0.53** | **0.23** | **-0.45** |
| AZE | **0.23** | **0.23** | 1 | 0.09 | -0.03 | -0.14 |
| RUS | **0.67** | **0.53** | 0.09 | 1 | -0.03 | **-0.27** |
| TUR | 0.01 | **0.23** | -0.03 | -0.03 | 1 | 0.17 |
| WRL | -0.14 | **-0.45** | -0.14 | **-0.27** | 0.17 | 1 |

## 3. Results and Discussion

The results in the Fig. 1-24 and Table 1-9 are presented.

**Fig. 1.**
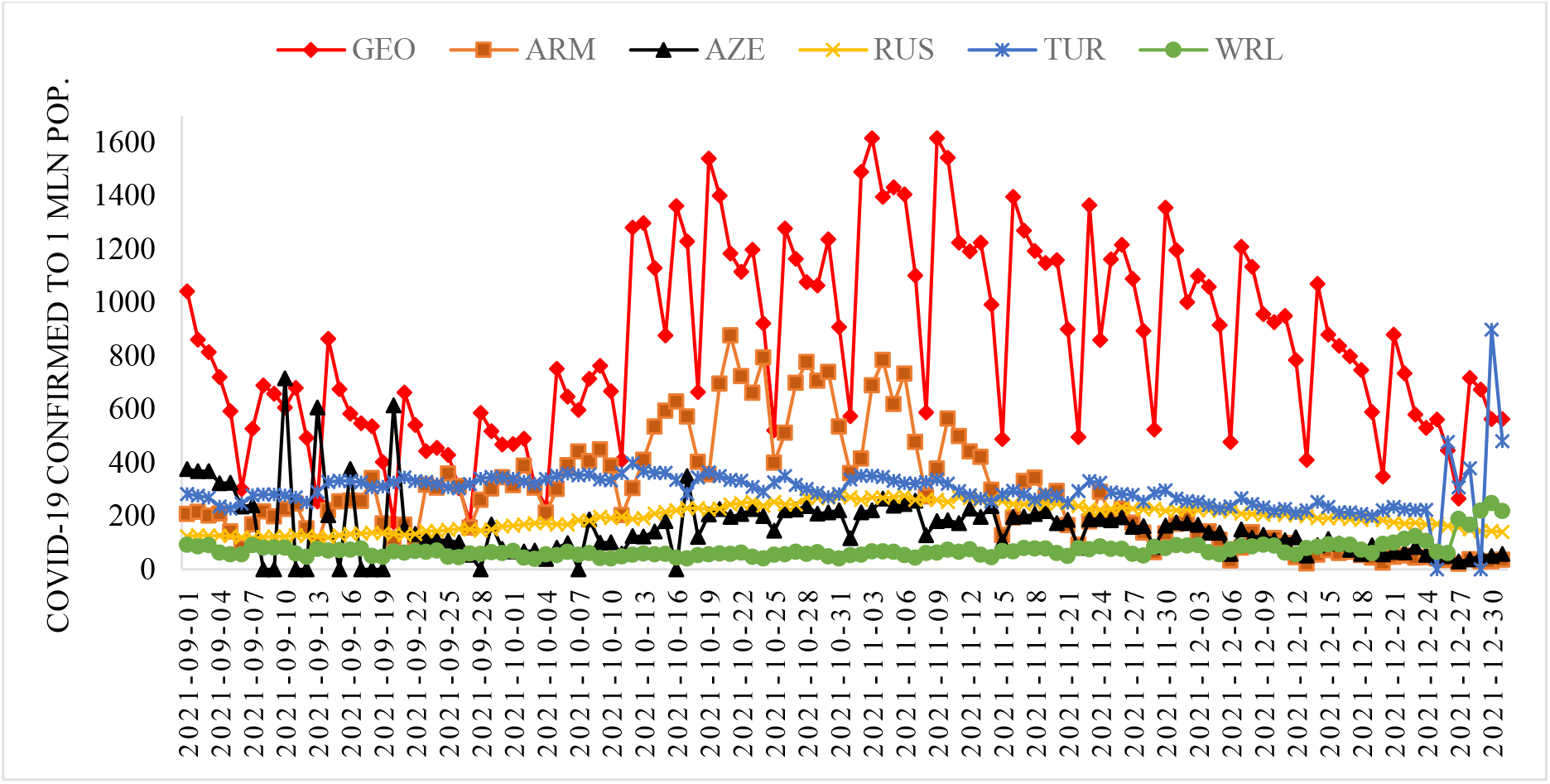
Time-series of Covid-19 confirmed cases to 1 million populations in Georgia, neighboring countries (Armenia, Azerbaijan, Russia, Turkey) and World from September 1, 2021 to December 31, 2021.

**Fig. 2.**
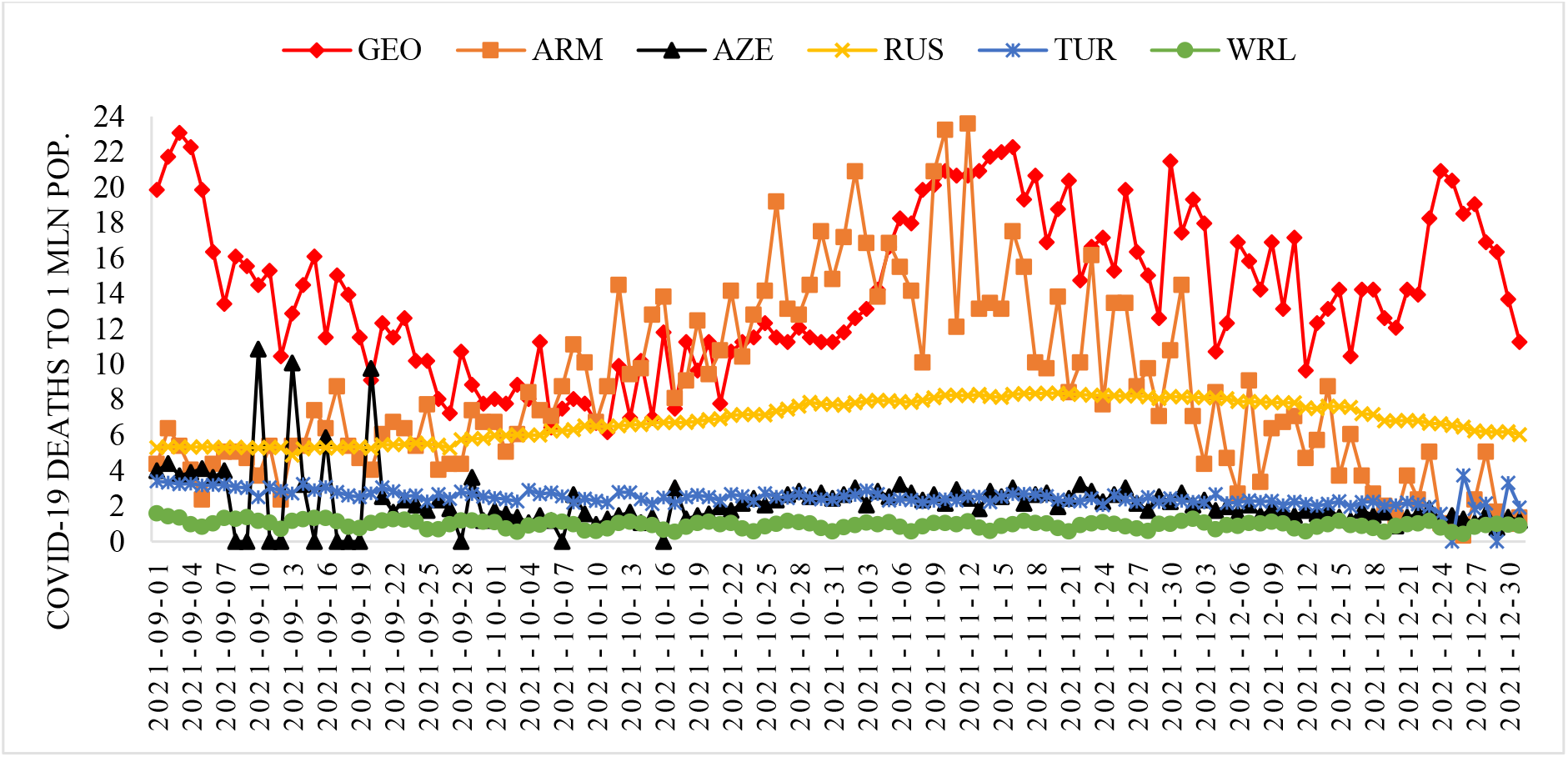
Time-series of deaths cases from Covid-19 to 1 million population in in Georgia, neighboring countries (Armenia, Azerbaijan, Russia, Turkey) and World from September 1, 2021 to December 31, 2021.

**Fig. 3.**
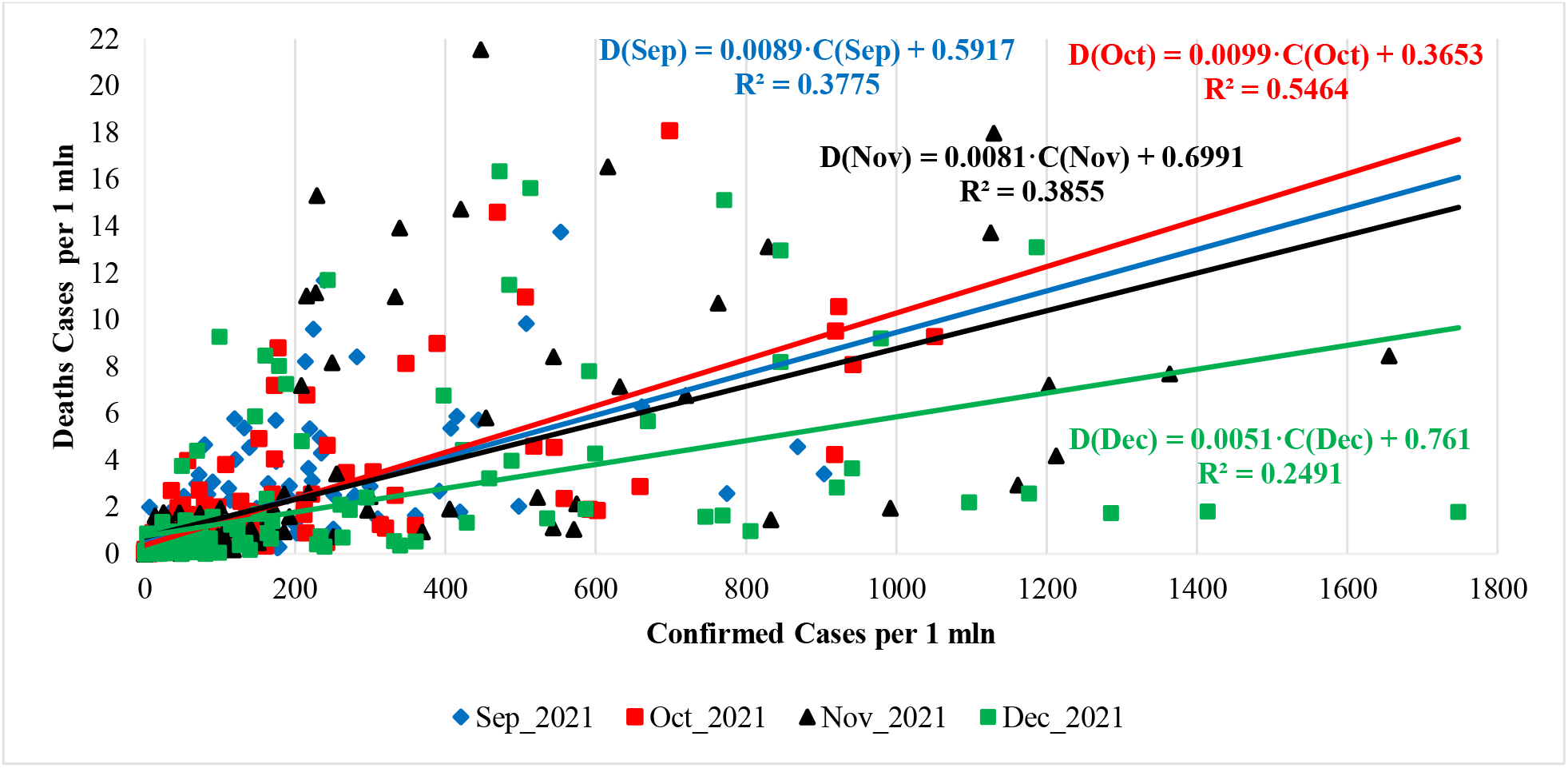
Linear correlation and regression between mean monthly values of deaths and confirmed cases related to Covid-19 for 157 countries with population ≥ 1 million inhabitants (normed on 1 mln pop.) from September to December 2021.

**Fig. 4.**
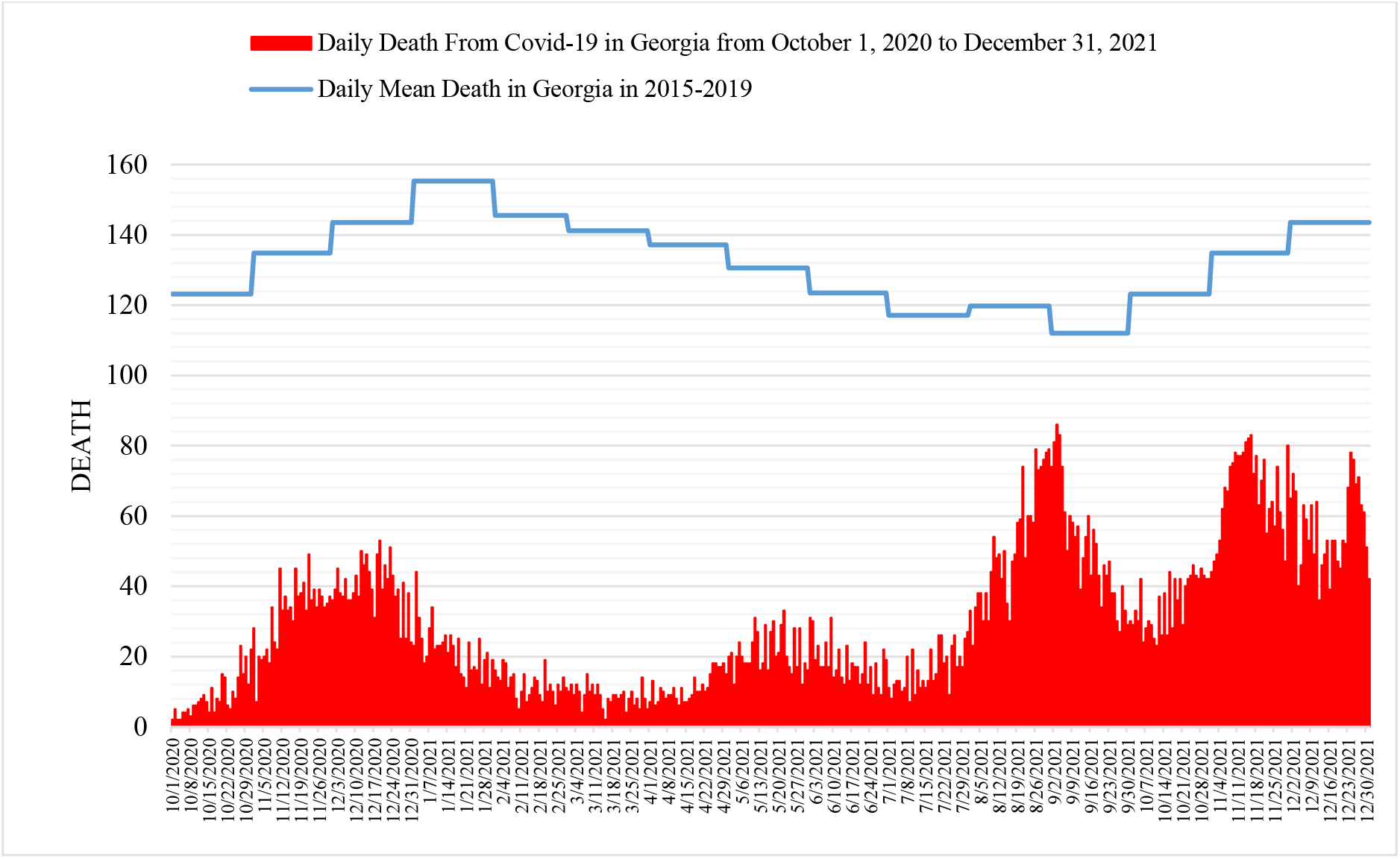
Daily death from Covid-19 in Georgia from September 1, 2020 to December 31, 2021 in comparison with daily mean death in 2015-2019.

**Fig. 5.**
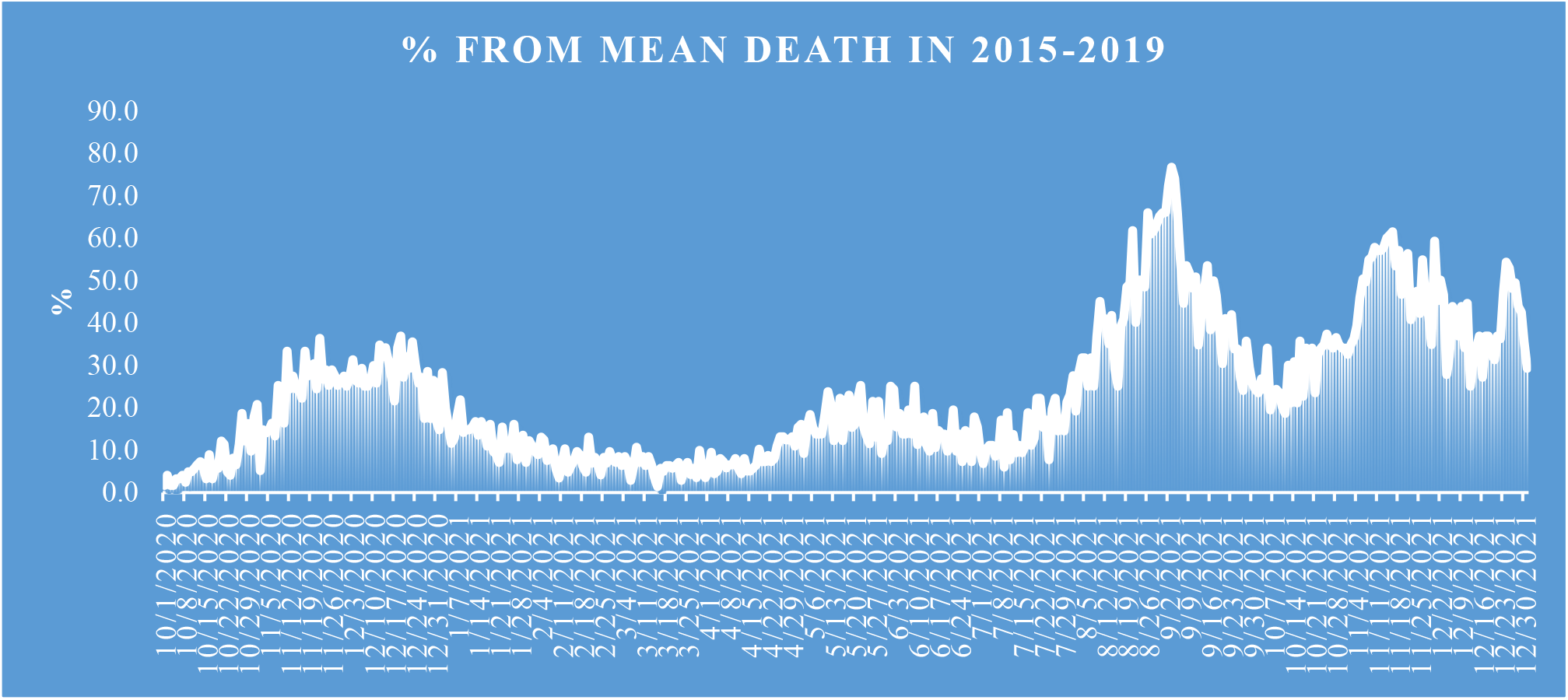
Share value of D from mean death in 2015-2019.

**Fig. 6.**
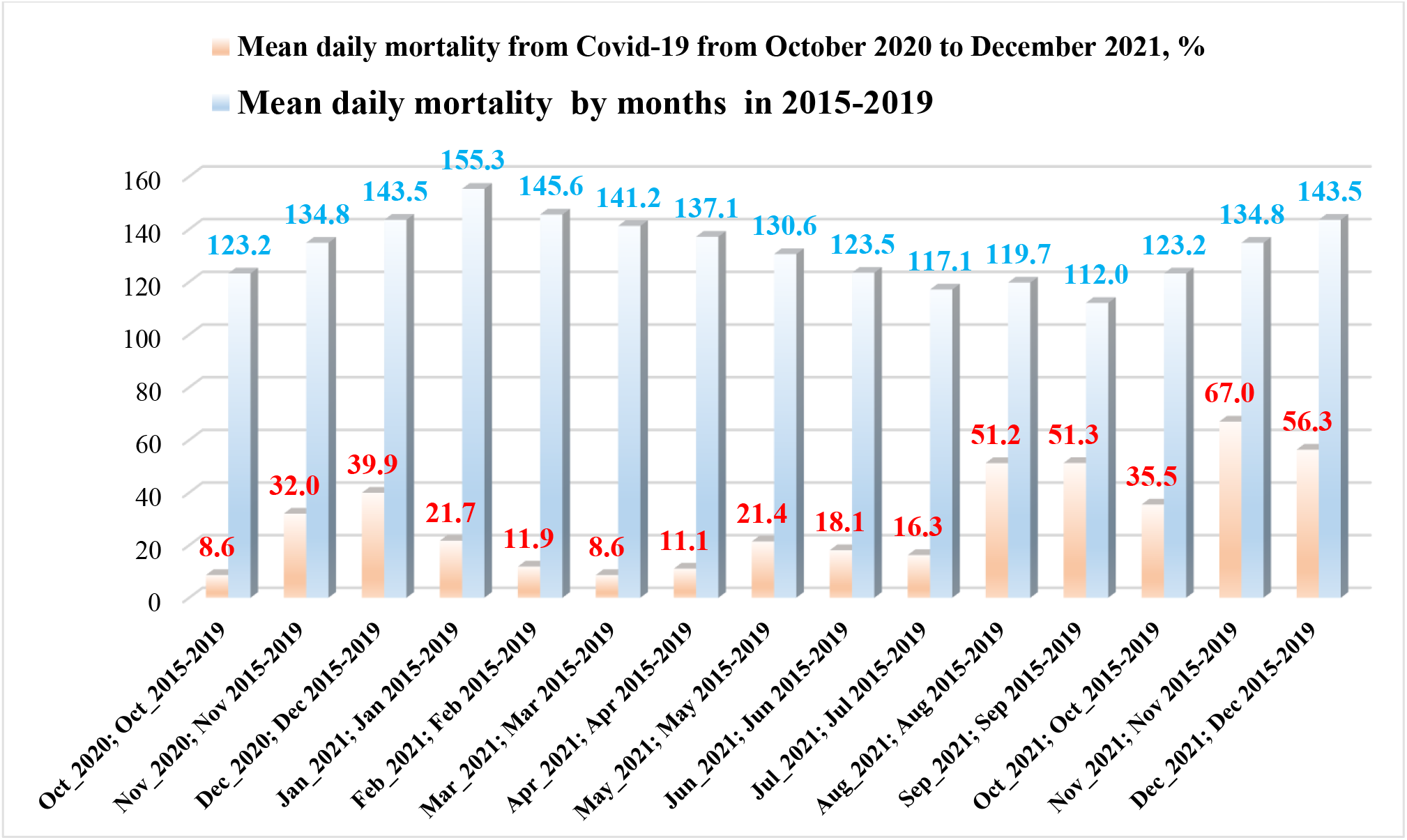
Mean daily mortality from Covid-19 from October 2020 to December 2021, % and mean daily mortality from January to December in 2015-2019.

**Fig. 7.**
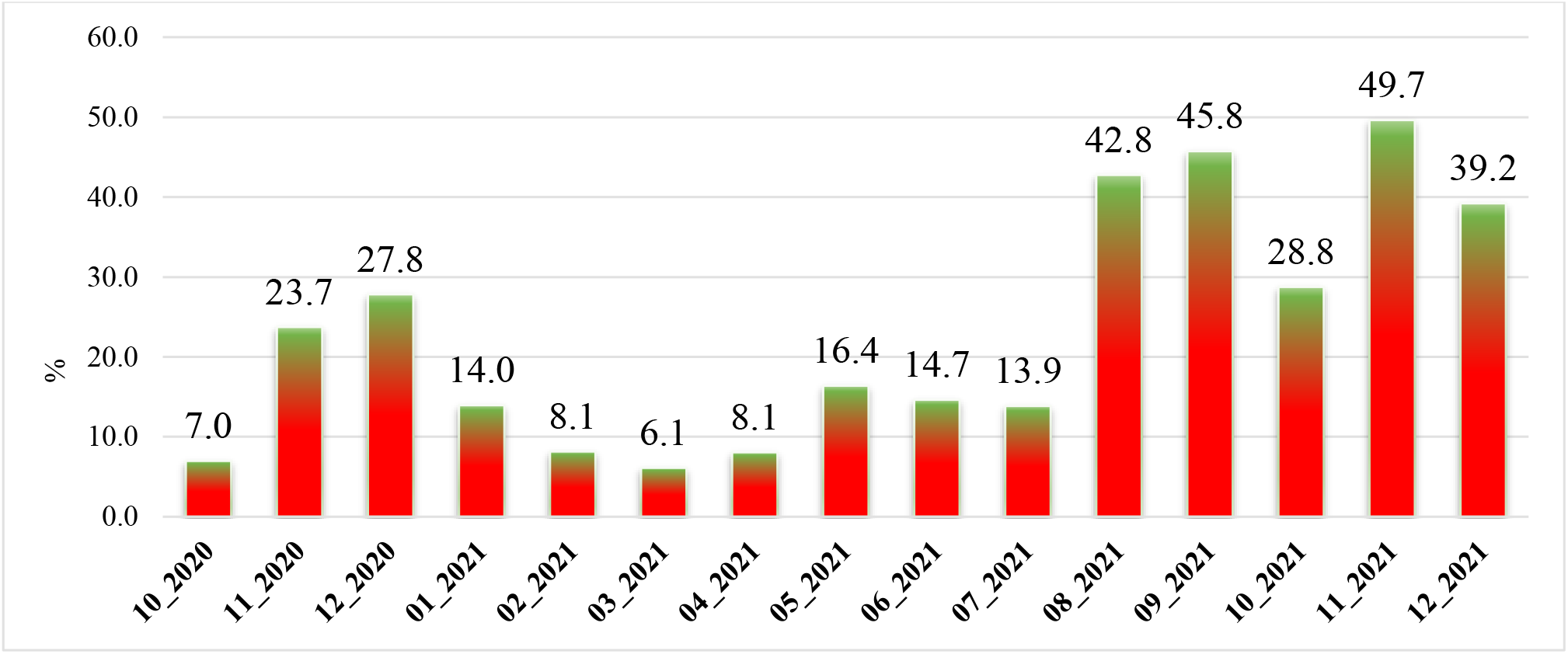
Share of mean daily mortality from Covid-19 of mean daily mortality in 2015-2019 from October 2020 to December 2021.

**Fig. 8.**
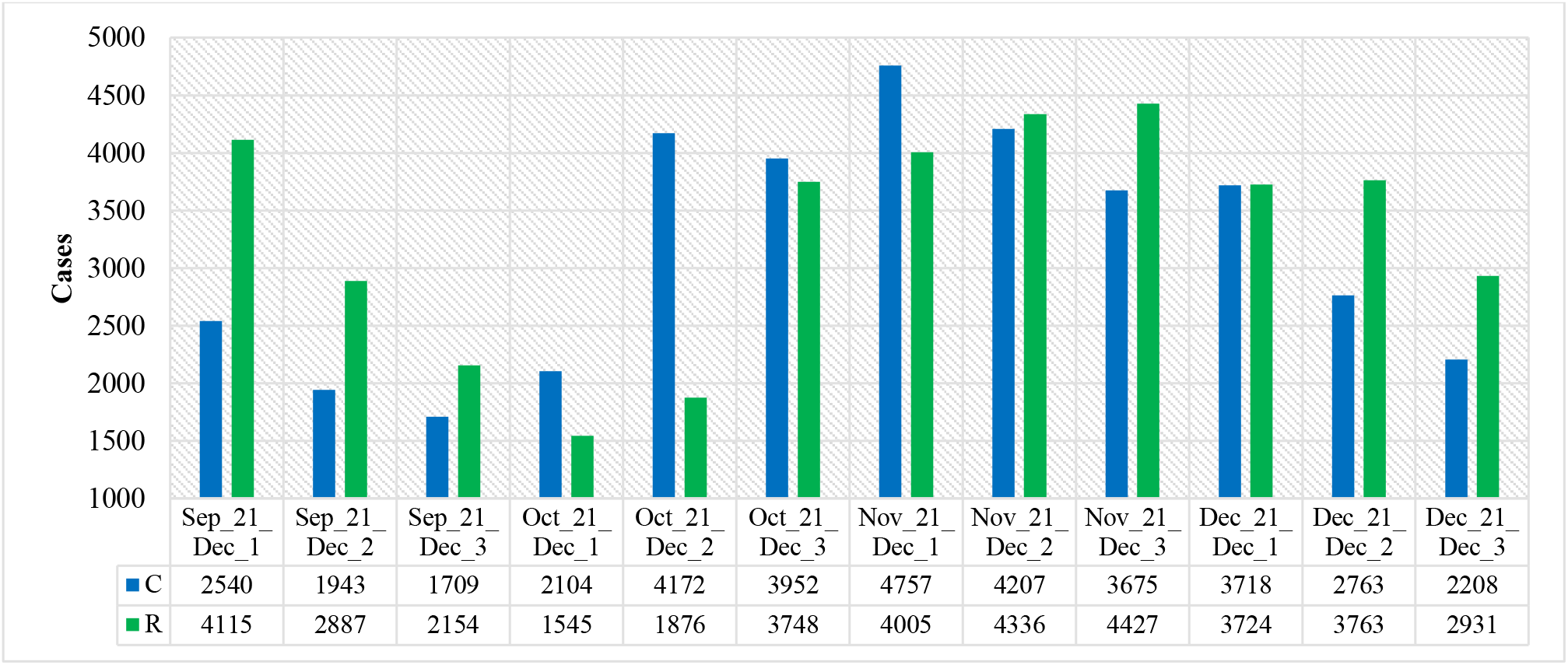
Mean values of confirmed and recovered coronavirus-related cases in different decades of months in Georgia from September 2021 to December 2021.

**Fig. 9.**
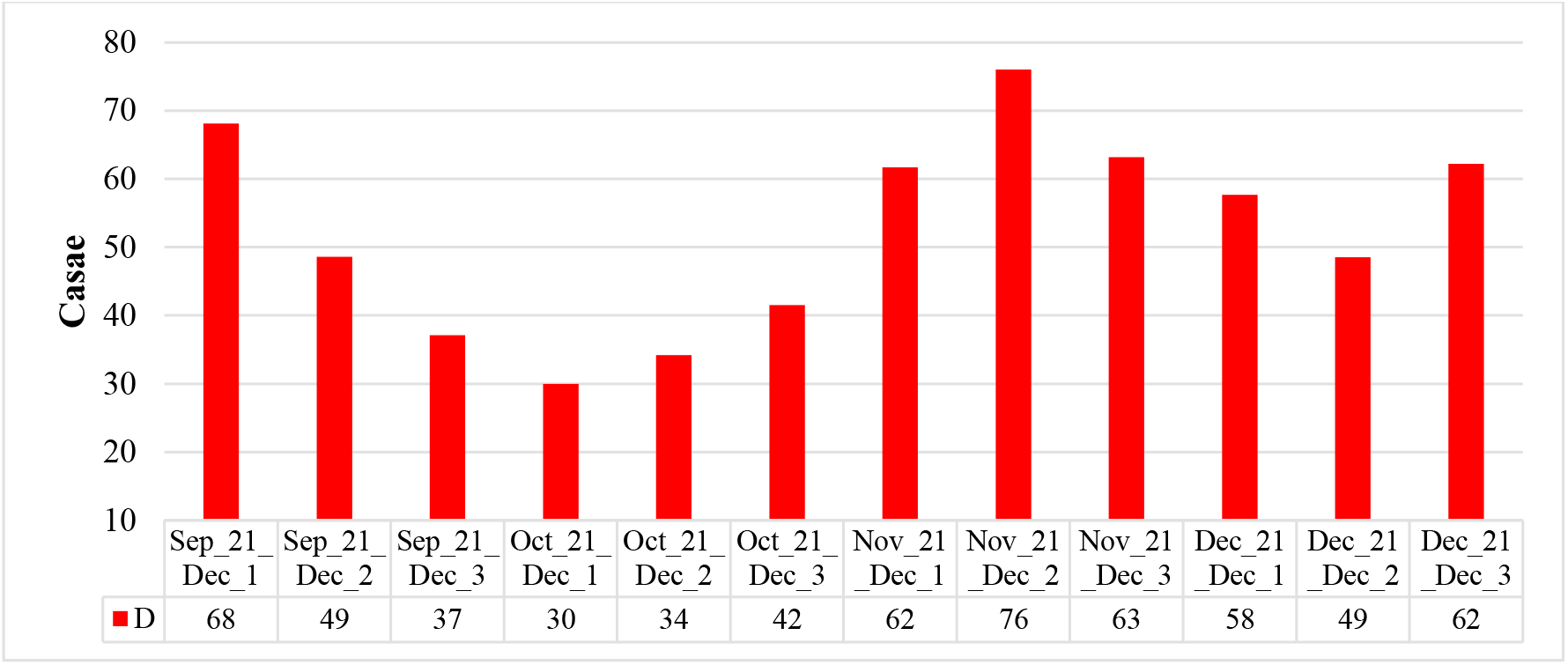
Mean values of deaths coronavirus-related cases in different decades of months in Georgia from September 2021 to December 2021.

**Fig. 10.**
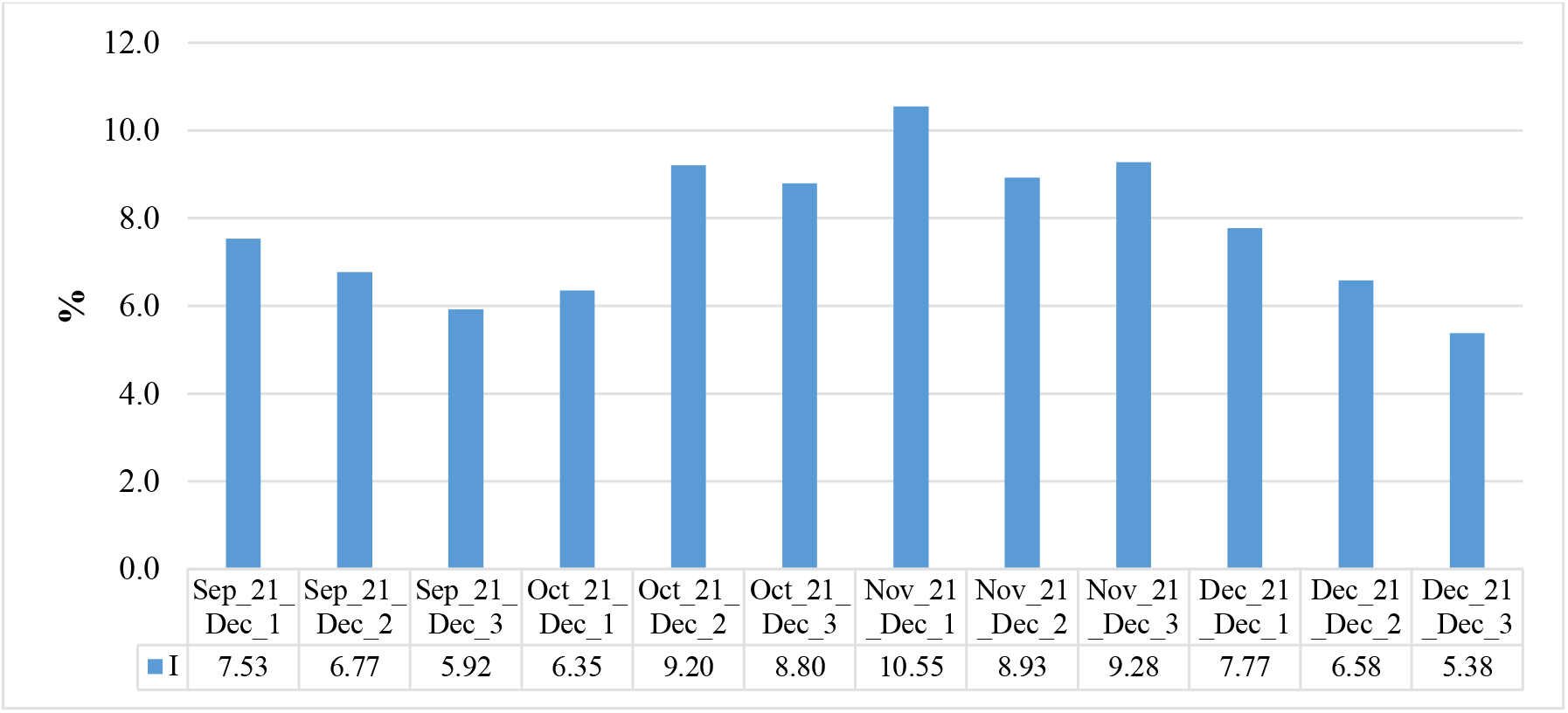
Mean values of infection rate coronavirus-related cases in different decades of months in Georgia from September 2021 to December 2021.

**Fig. 11.**
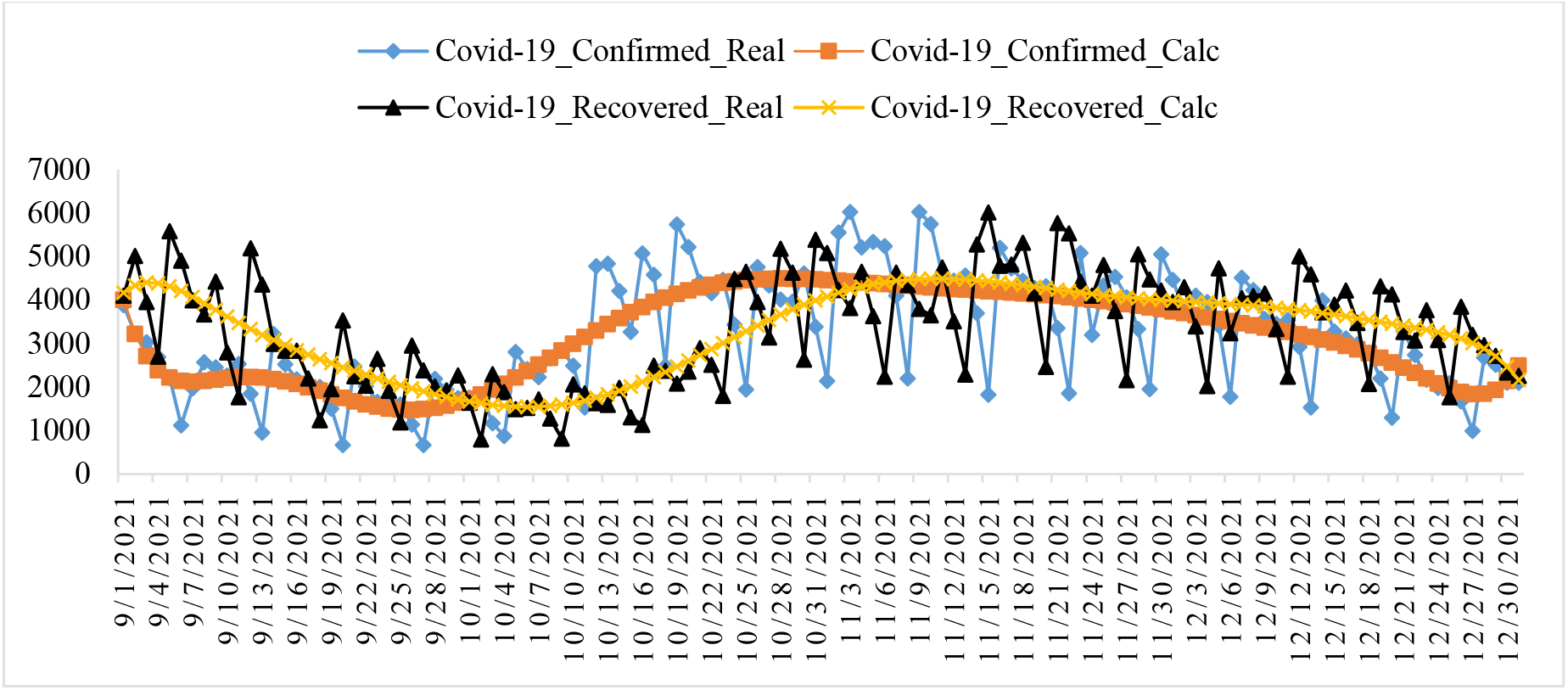
Changeability of the real and calculated daily values of coronavirus confirmed and recovered cases from September 01, 2021 to December 31, 2021 in Georgia.

**Fig. 12.**
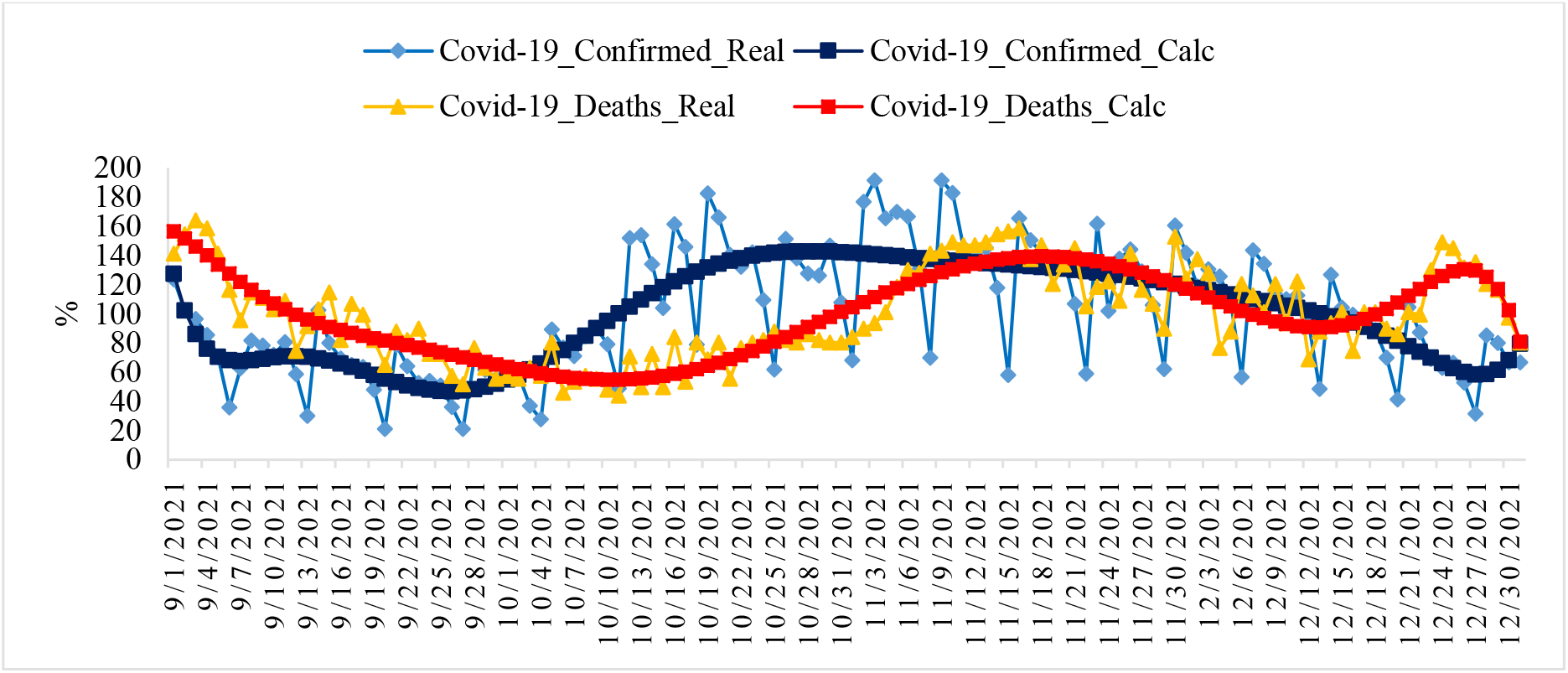
Changeability of the real and calculated daily values of coronavirus-related confirmed and deaths cases from September 01, 2021 to December 31, 2021 in Georgia (normed on mean values, %).

**Fig. 13.**
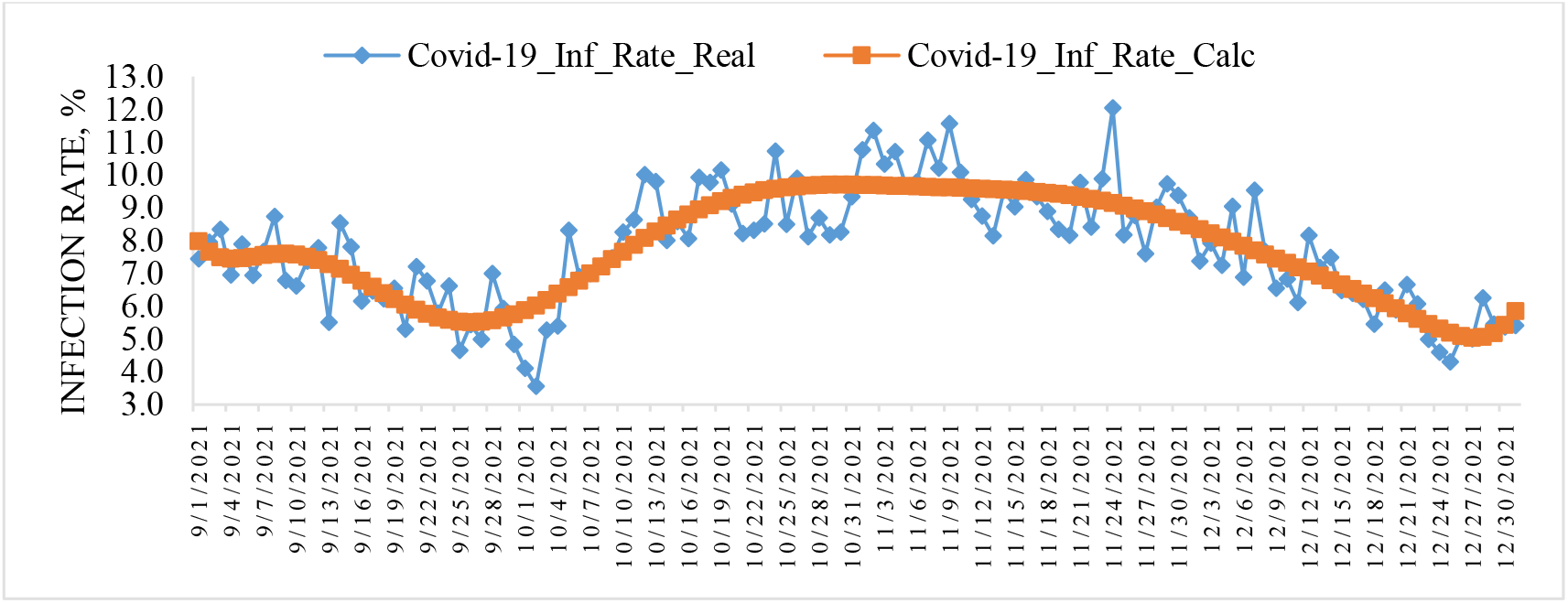
Changeability of the real and calculated daily values of coronavirus Infection Rate from September 01, 2021 to December 31, 2021 in Georgia.

**Fig. 14.**
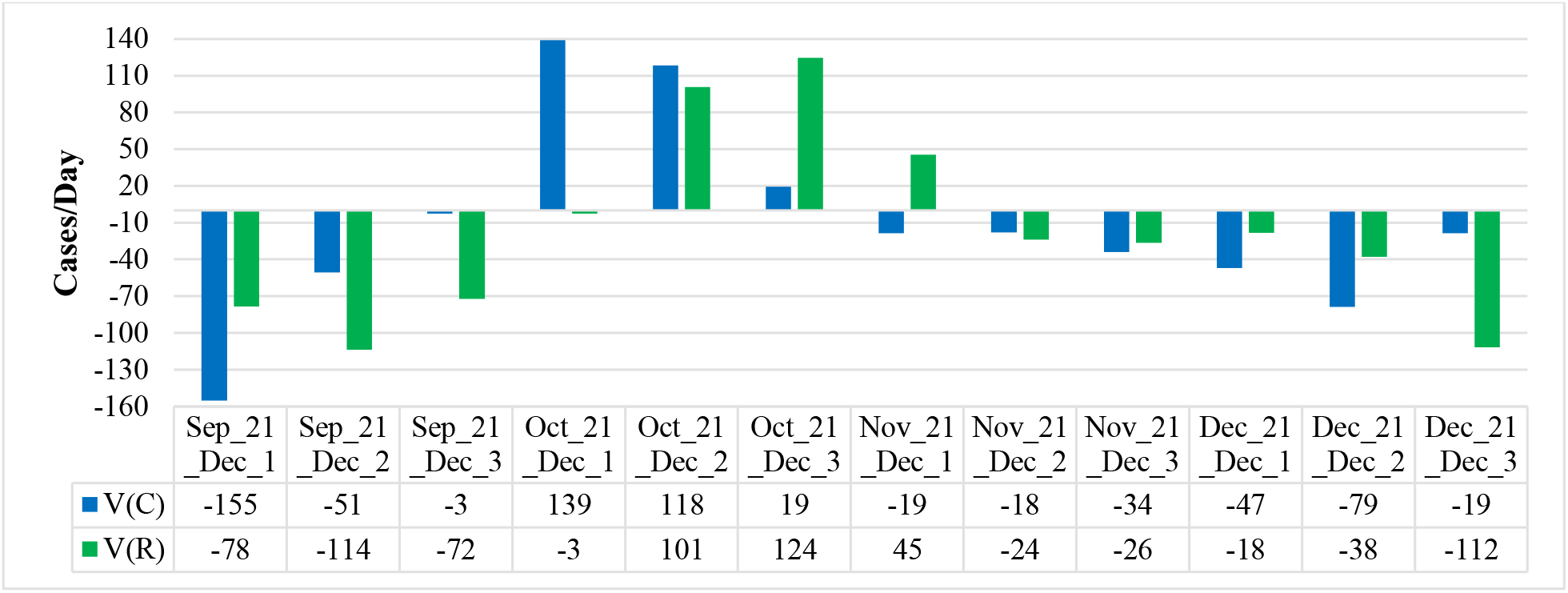
Mean values of speed of change of confirmed and recovered coronavirus-related cases in different decades of months in Georgia from September 2021 to December 2021.

**Fig. 15.**
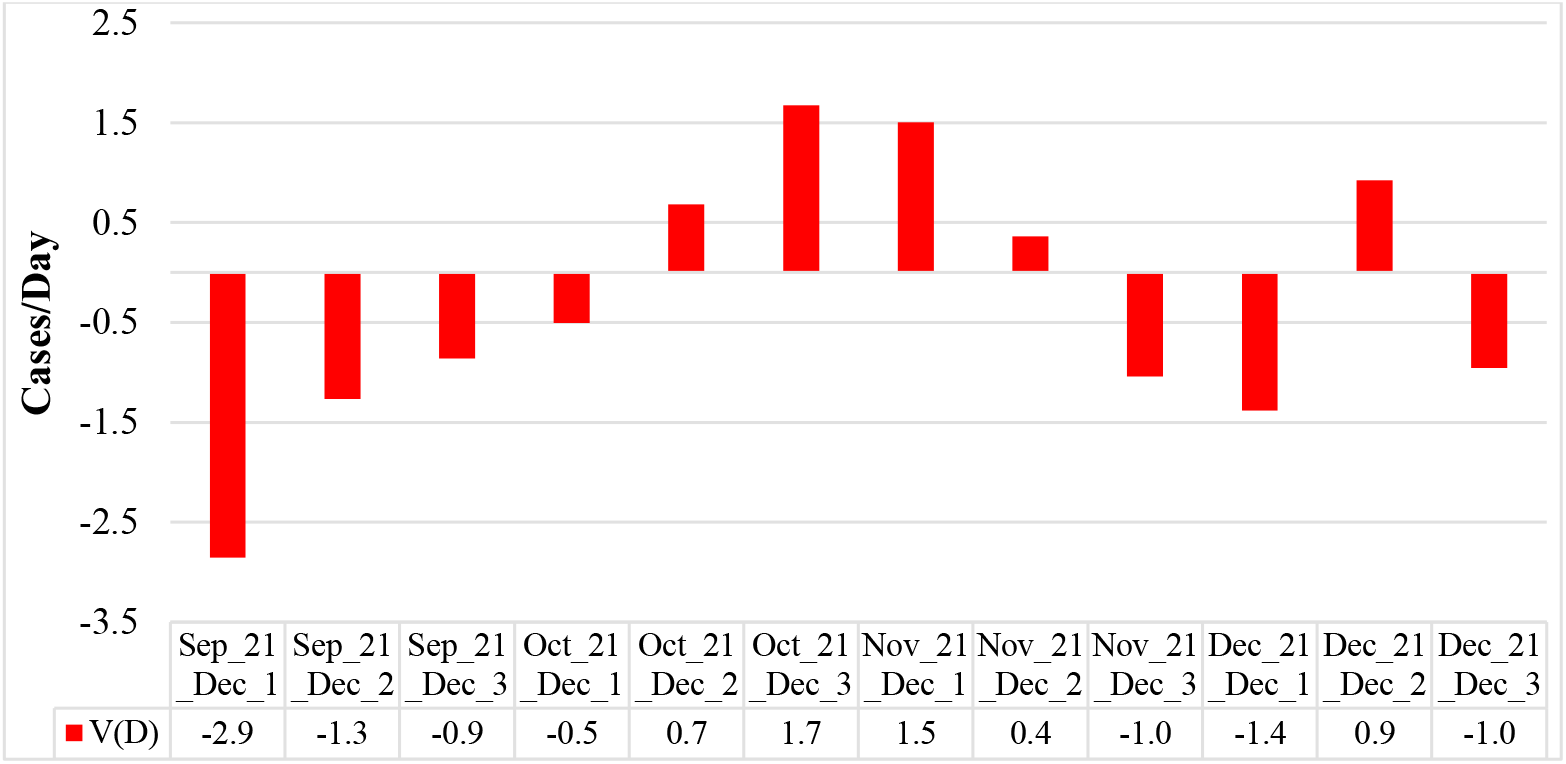
Mean values of speed of change of deaths coronavirus-related cases in different decades of months in Georgia from September 2021 to December 2021.

**Fig. 16.**
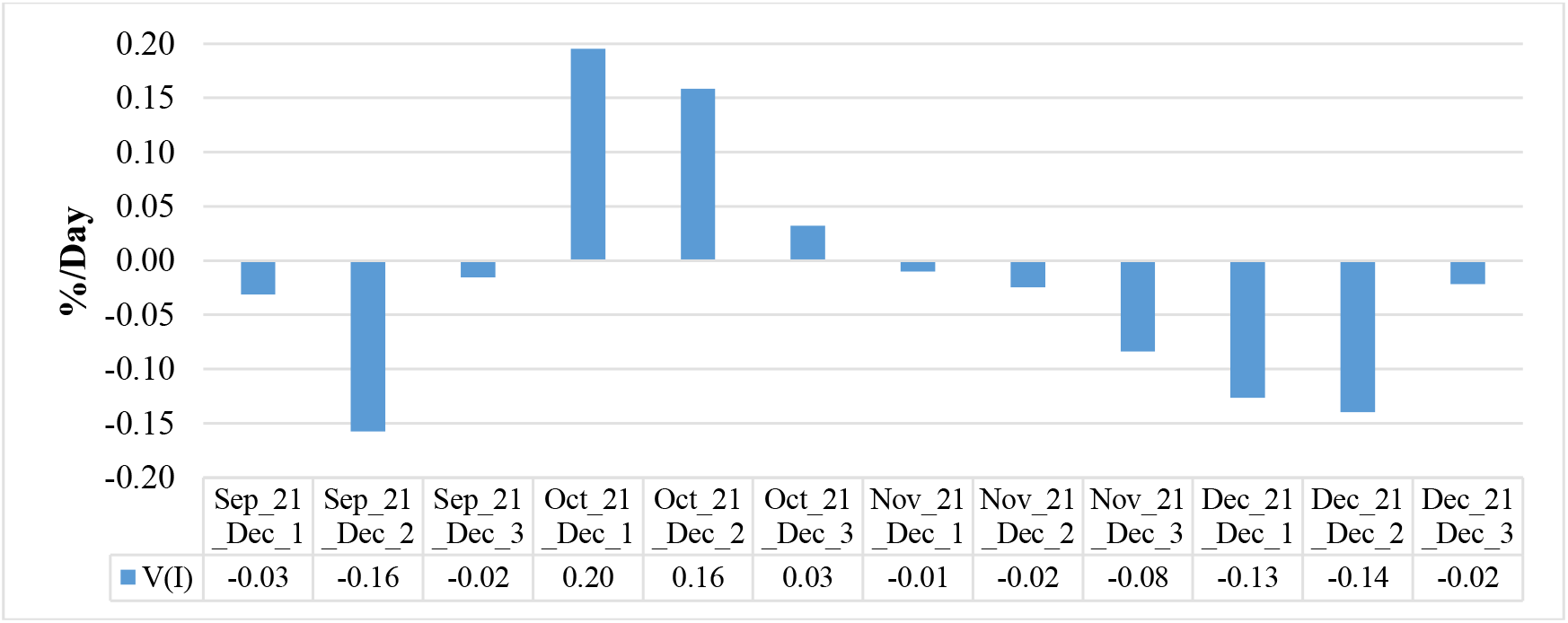
Mean values of speed of change of coronavirus infection rate in different decades of months in Georgia from September 2021 to December 2021.

**Fig. 17.**
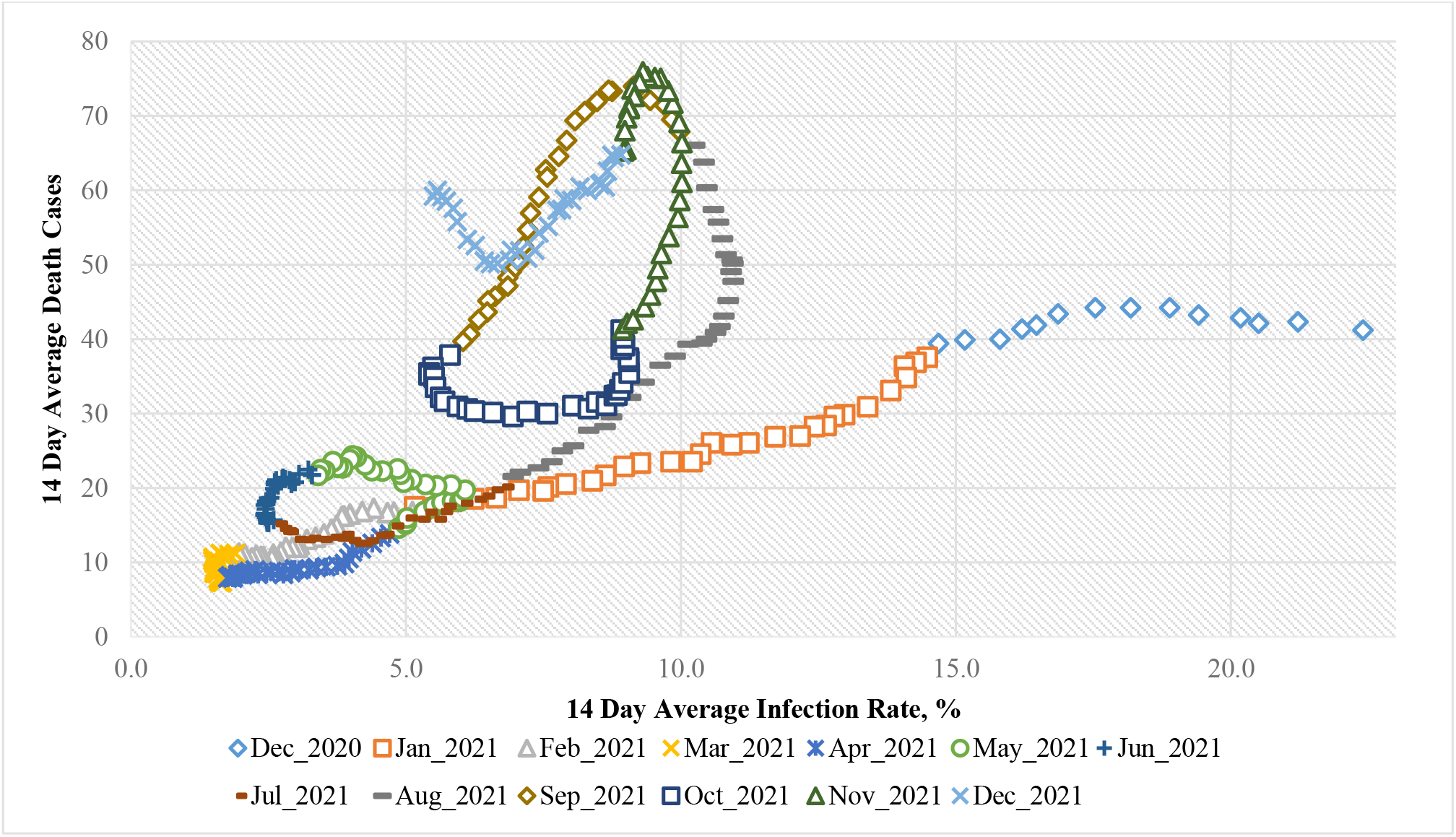
Connection of 14-day moving average of deaths cases due to COVID-19 in Georgia with 14-day moving average of infection rate from December 18, 2020 until December 31, 2021.

**Fig. 18.**
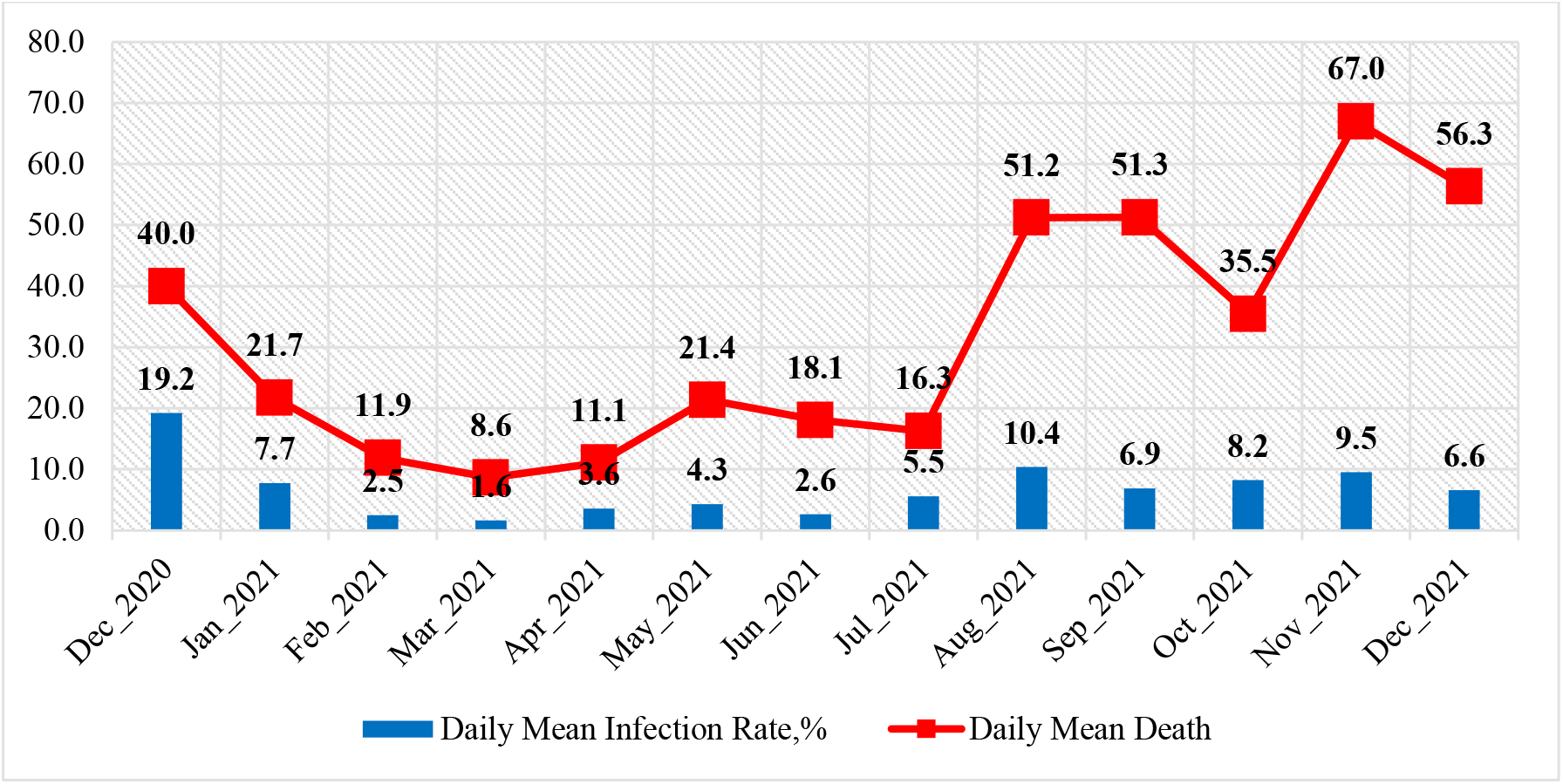
Mean monthly values of COVID-19 infection rate and deaths cases in Georgia from December 2020 to December 2021.

**Fig. 19.**
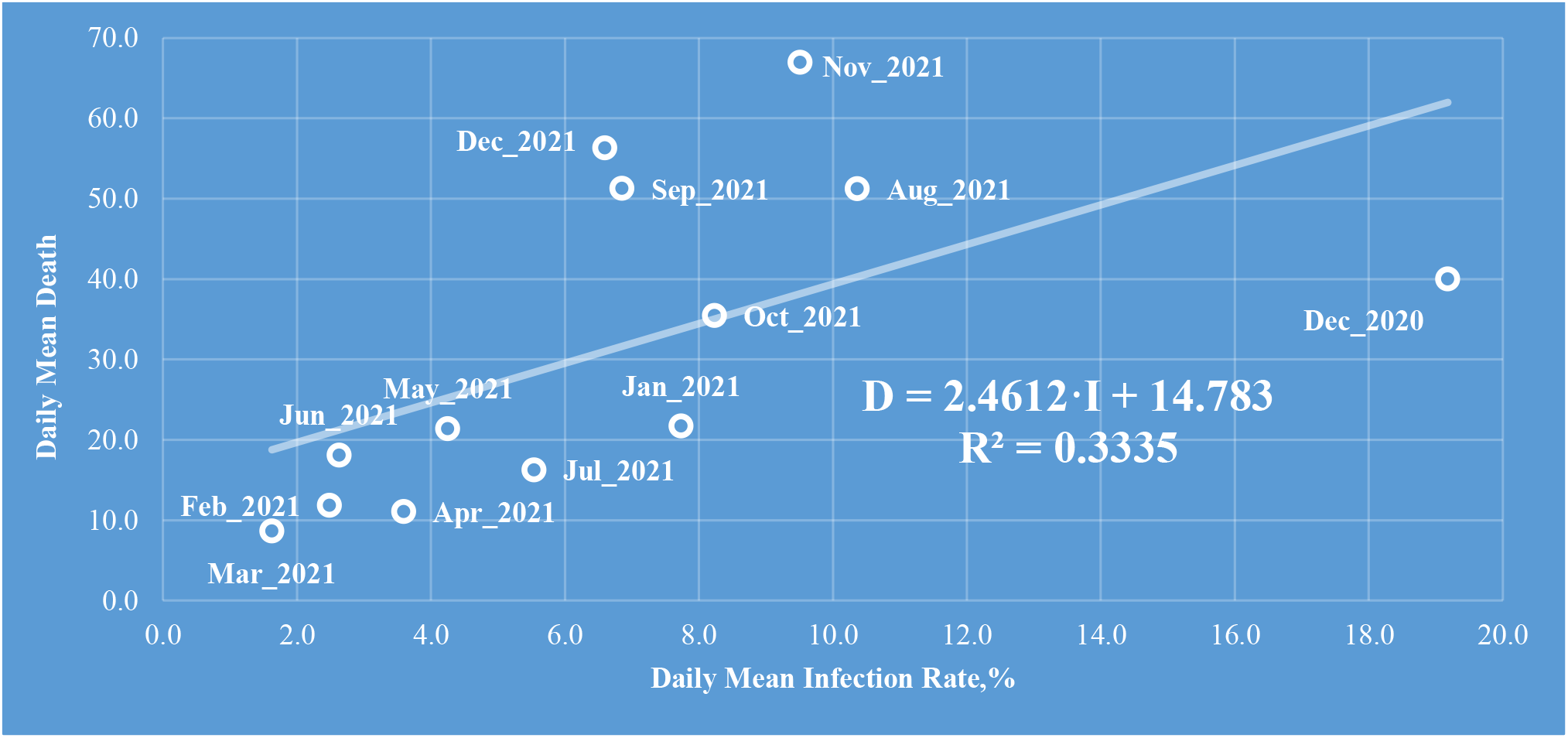
Linear correlation and regression between the mean monthly values of COVID-19 infection rate and deaths cases in Georgia from December 2020 to December 2021. R=0.58 (moderate correlation [41]), α≈0.03

**Fig. 20.**
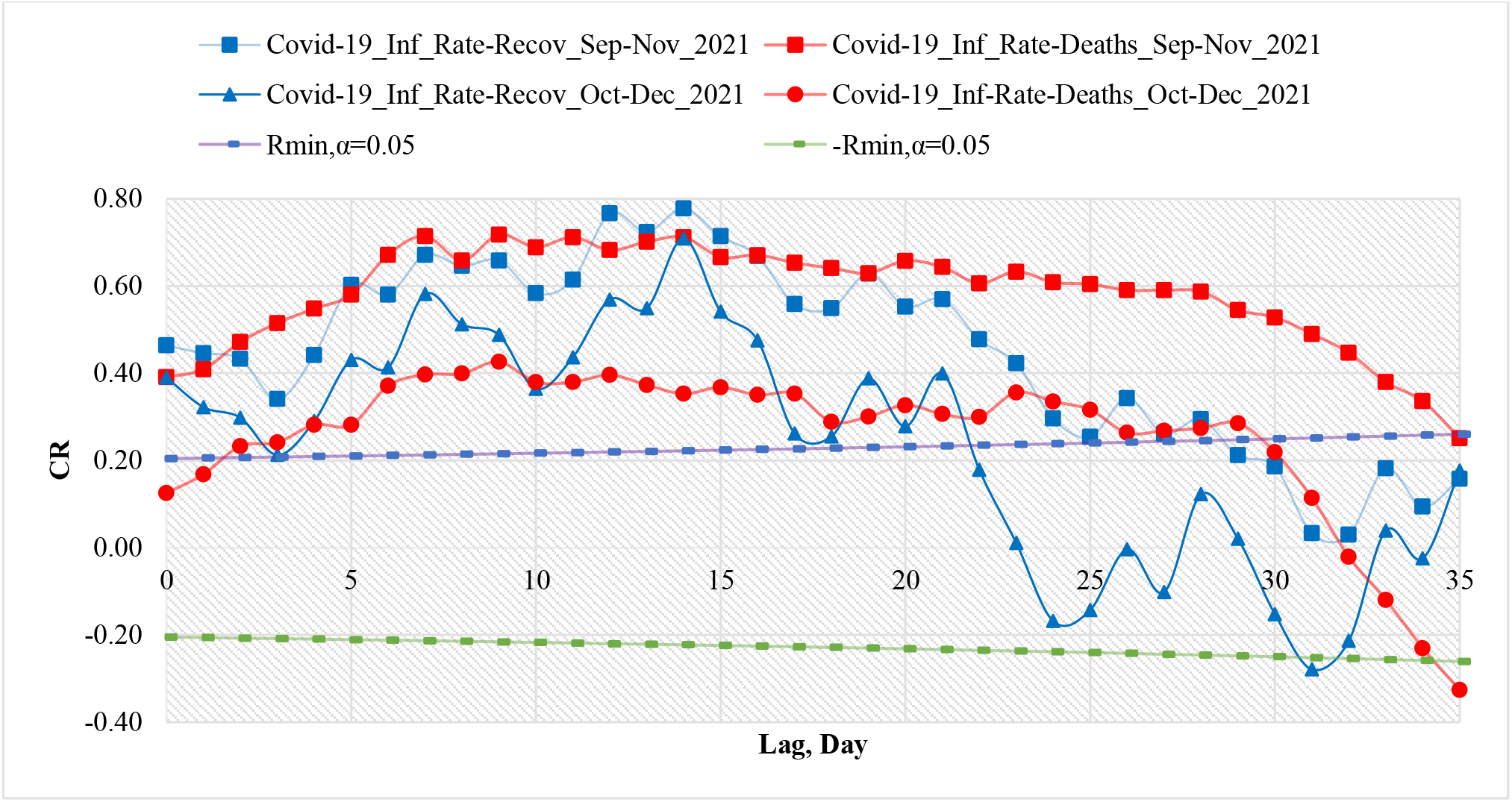
Coefficient of cross-correlations between confirmed COVID-19 cases (normed to tests number) with recovered and deaths cases in Georgia from 01.09.2021 to 30.11.2021 and from 01.10.2021 to 31.12.2021.

**Fig. 21.**
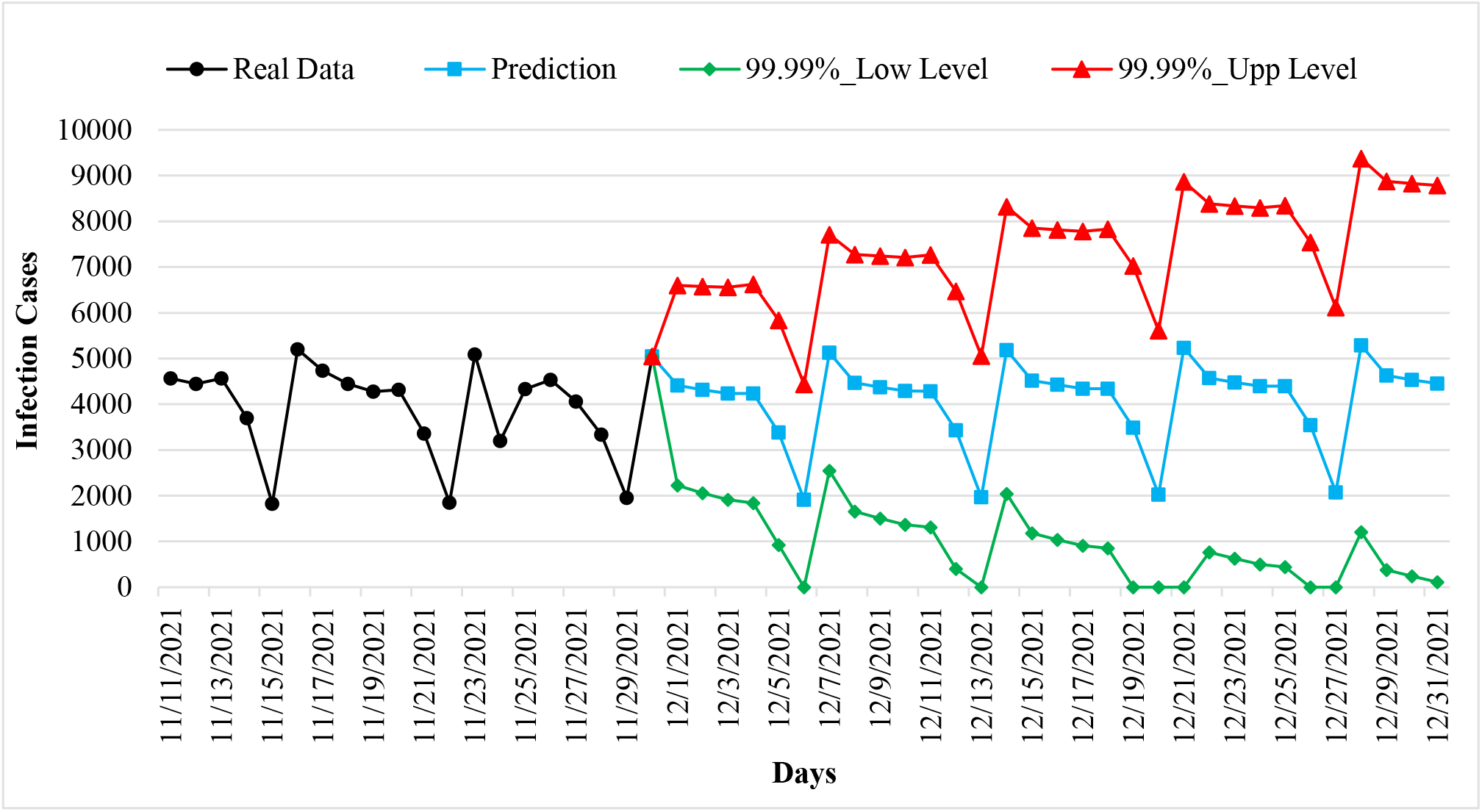
Example of Interval Prediction of COVID-19 Infection Cases in Georgia from 01.12.2021 to 31.12.2021.

**Fig. 22.**
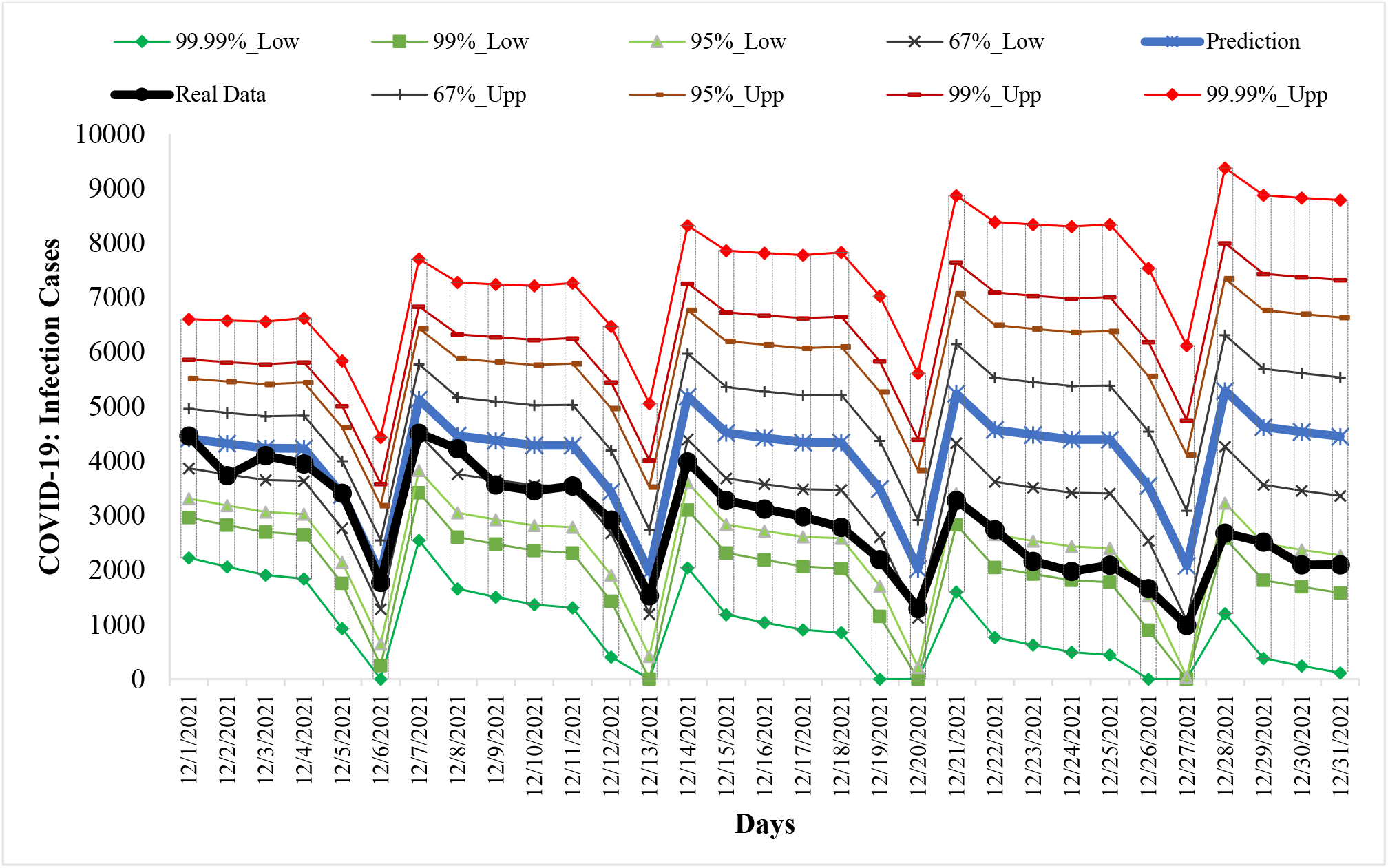
Example of Verification of Interval Prediction of COVID-19 Infection Cases in Georgia from 01.12.2021 to 31.12.2021.

**Fig. 23.**
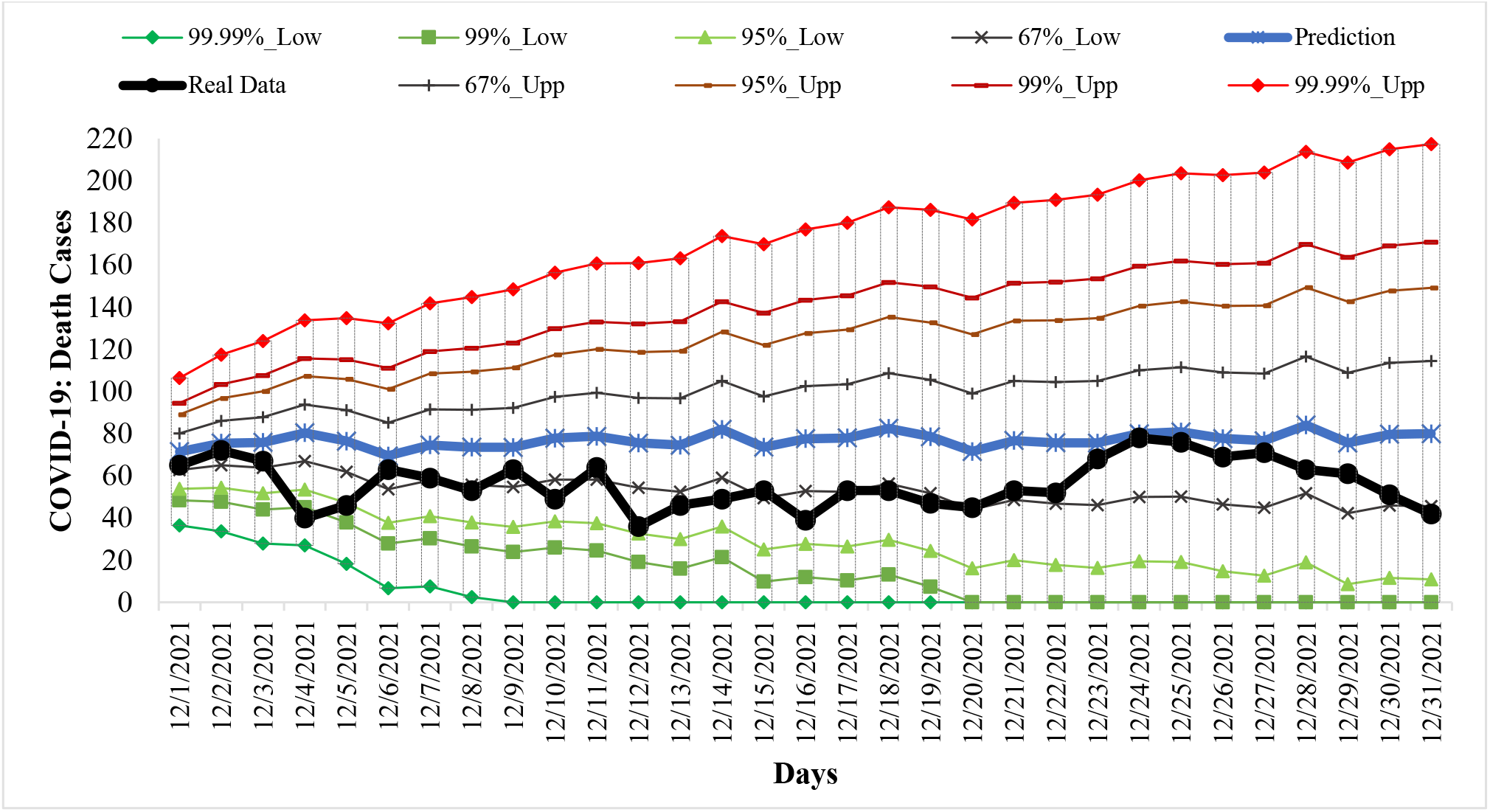
Example of Verification Interval Prediction of COVID-19 Death Cases in Georgia from 01.12.2021 to 31.12.2021.

**Fig. 24.**
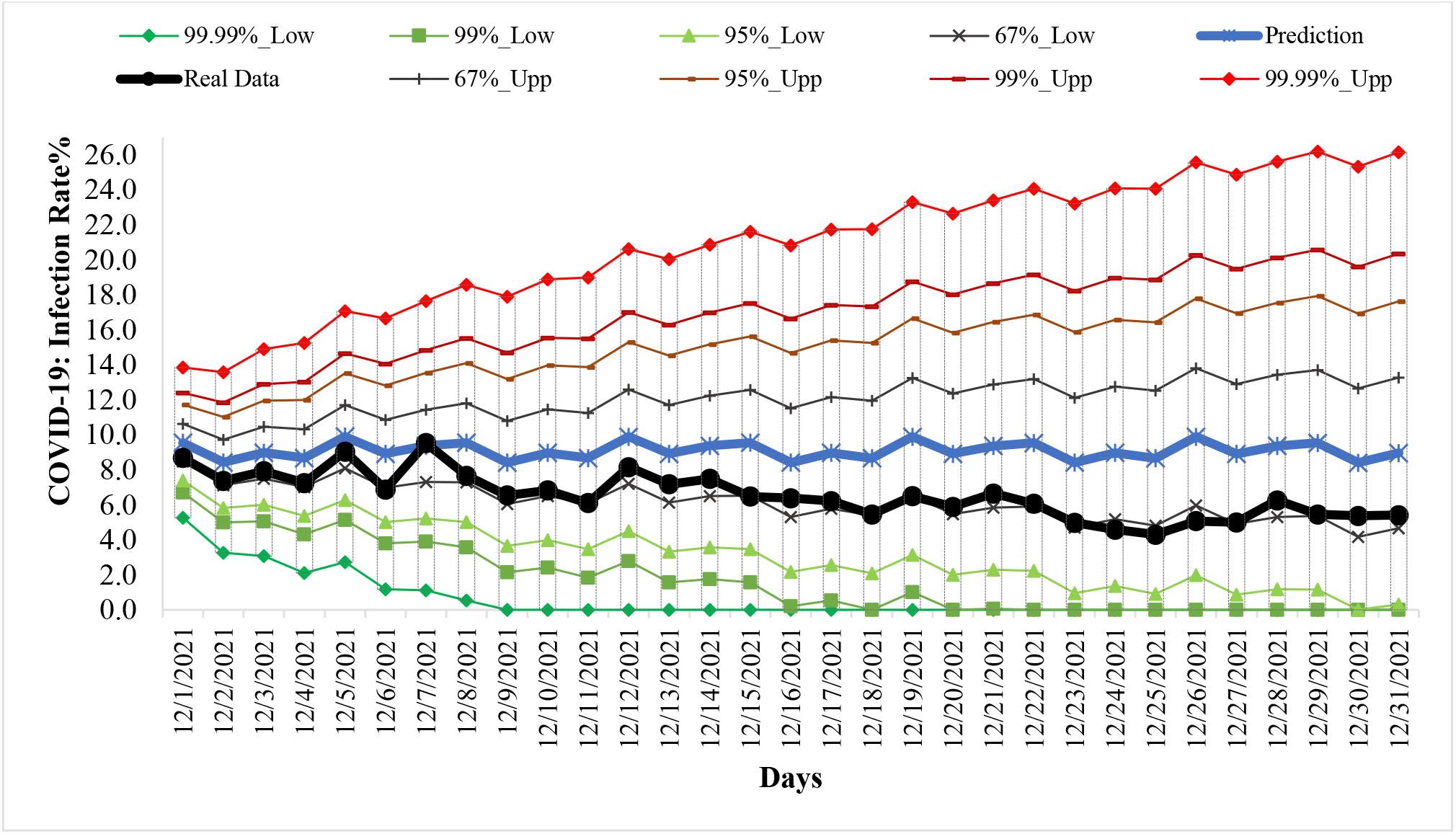
Example of Verification Interval Prediction of COVID-19 Infection Rate in Georgia from 01.12.2021 to 31.12.2021.

**Table 2.**
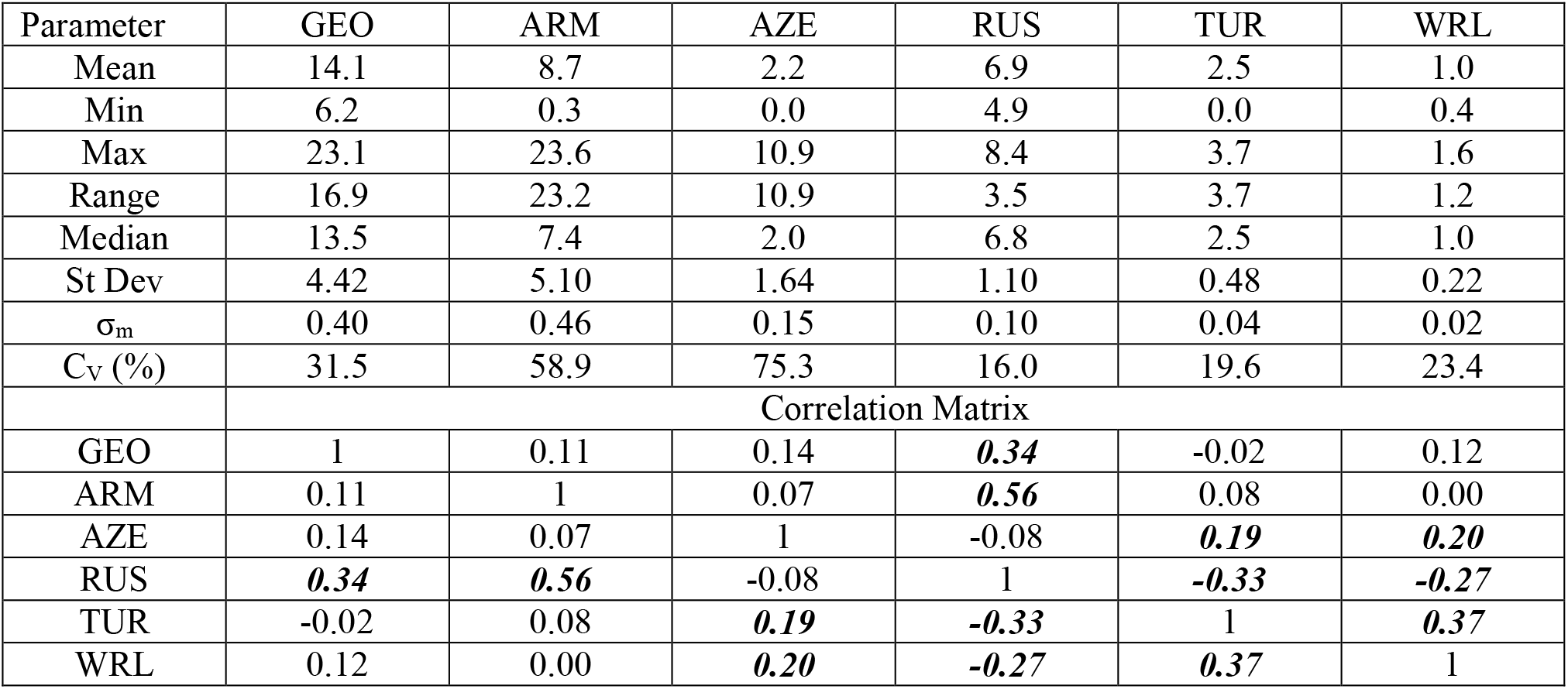
The statistical of deaths cases from Covid-19 to 1 million populations in Georgia, neighboring countries (Armenia, Azerbaijan, Russia, Turkey) and World from September 1, 2021 to December 31, 2021. (r_min_ = ± 0.18, α = 0.05).

**Table 3.**
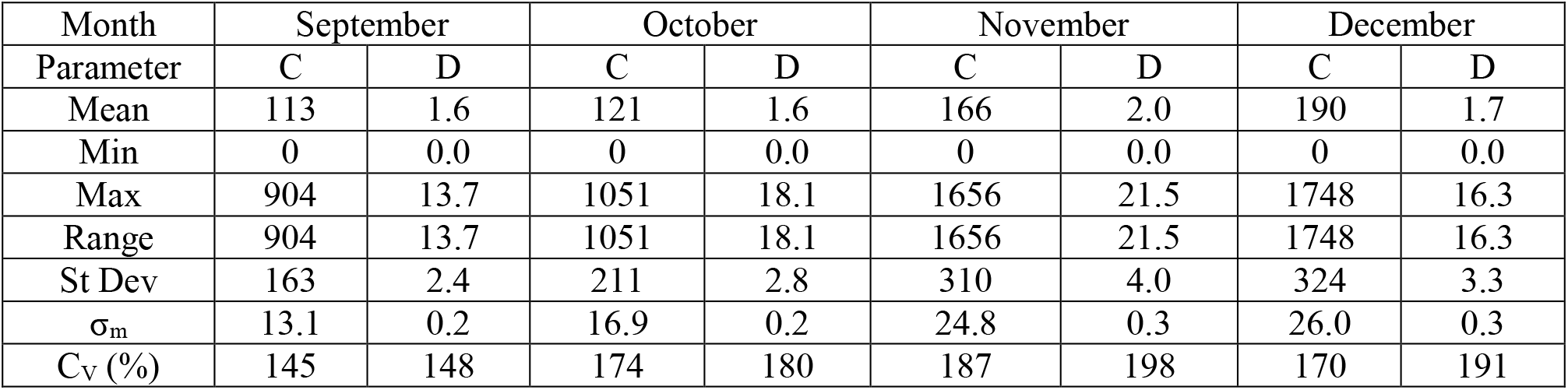
The statistical characteristics of mean monthly values of confirmed cases and deaths cases related to Covid-19 for 157 countries with population ≥ 1 million inhabitant from September to December 2021 (normed on 1 mln pop.).

| Month | September |  | October |  | November |  | December |  |
| --- | --- | --- | --- | --- | --- | --- | --- | --- |
| Parameter | C | D | C | D | C | D | C | D |
| Mean | 113 | 1.6 | 121 | 1.6 | 166 | 2.0 | 190 | 1.7 |
| Min | 0 | 0.0 | 0 | 0.0 | 0 | 0.0 | 0 | 0.0 |
| Max | 904 | 13.7 | 1051 | 18.1 | 1656 | 21.5 | 1748 | 16.3 |
| Range | 904 | 13.7 | 1051 | 18.1 | 1656 | 21.5 | 1748 | 16.3 |
| St Dev | 163 | 2.4 | 211 | 2.8 | 310 | 4.0 | 324 | 3.3 |
| $\sigma_m$ | 13.1 | 0.2 | 16.9 | 0.2 | 24.8 | 0.3 | 26.0 | 0.3 |
| $C_V$ (%) | 145 | 148 | 174 | 180 | 187 | 198 | 170 | 191 |

**Table 4.** Covid-19 mean monthly values of infection (C) and deaths (D) cases from September to December 2021 (per 1 million population) and ranking of Georgia, Armenia, Azerbaijan, Russia, Turkey by these parameters (in brackets) among 157 countries with population ≥ 1 million inhabitant, and correspondent values of deaths coefficient (DC).

| Location | Georgia | Armenia | Azerbaijan | Russia | Turkey | World |
| --- | --- | --- | --- | --- | --- | --- |
| Parameter/Month | September |  |  |  |  |  |
| C (Rating C) | 553.5 (5) | 219.7 (27) | 192.4 (32) | 132.6 (44) | 300.1 (15) | 67.0 |
| D ( Rating D) | 13.75 (1) | 5.33 (14) | 2.90 (29) | 5.38 (12) | 2.88 (30) | 1.12 |
| DC, % | 2.48 | 2.43 | 1.51 | 4.06 | 0.96 | 1.67 |
| Parameter/Month | October |  |  |  |  |  |
| C (Rating C) | 919.1 (4) | 506.8 (13) | 149.8 (37) | 216.0 (26) | 333.4 (18) | 53.1 |
| D ( Rating D) | 9.51 (5) | 10.97 (3) | 1.73 (39) | 6.77 (12) | 2.49 (28) | 0.89 |
| DC, % | 1.03 | 2.16 | 1.16 | 3.14 | 0.75 | 1.67 |
| Parameter/Month | November |  |  |  |  |  |
| C (Rating C) | 1130.0 (6) | 339.1 (25) | 185.7 (38) | 249.2 (31) | 299.7 (27) | 66.2 |
| D ( Rating D) | 17.96 (2) | 13.91 (6) | 2.55 (26) | 8.16 (15) | 2.44 (27) | 0.91 |
| DC, % | 1.59 | 4.10 | 1.37 | 3.27 | 0.81 | 1.38 |
| Parameter/Month | December |  |  |  |  |  |
| C (Rating C) | 770.8 (13) | 69.7 (66) | 90.3 (58) | 188.3 (41) | 260.6 (35) | 104.0 |
| D ( Rating D) | 15.11 (3) | 4.46 (19) | 1.58 (39) | 7.24 (14) | 2.09 (31) | 0.90 |
| DC, % | 1.96 | 6.39 | 1.75 | 3.85 | 0.80 | 0.86 |

**Table 5.** Statistical characteristics of the daily data associated with coronavirus COVID-19 pandemic of confirmed, recovered, deaths cases and infection rate of the population of Georgia from 01.09.2021 to 31.12.2021.

| Parameter | Confirmed | Recovered | Deaths | Infection Rate, % |
| --- | --- | --- | --- | --- |
| Mean | 3145 | 3293 | 52 | 7.74 |
| Min | 662 (Sep 20) | 795 (Oct 2) | 23 (Oct 11) | 3.56 (Oct 2) |
| Max | 6024 (Nov 3) | 6017 (Nov 15) | 86 (Sep 3) | 12.04 (Nov 24) |
| Range | 5362 | 5222 | 63 | 8.48 |
| St Dev | 1336 | 1274 | 16 | 1.79 |
| $\sigma_m$ | 121 | 116 | 1.5 | 0.16 |
| $C_v$ (%) | 42.5 | 38.7 | 31.5 | 23.2 |

**Table 6.** Form of the equations of the regression of the time changeability of the daily values of C, R and D from September 01, 2021 to December 31, 2021 in Georgia. The level of significance of R^2^ is not worse than 0.001.

| Parameter | Confirmed | Recovered | Deaths | Infection Rate |
| --- | --- | --- | --- | --- |
| Regression | Tenth Order Polynomial |  |  |  |
| $R^2$ | 0.577 | 0.565 | 0.789 | 0.726 |
| $K_{DW}$ | 1.97 | 2.01 | 1.57 | 1.61 |
| Autocorrelation of residual components | Absent |  |  |  |

**Table 7.** Mean monthly values of C, R, D, I and its speed of change in Georgia from September 2021 to December 2021.

| Parameter | C | R | D | I, % | V(C) | V(R) | V(D) | V(I) |
| --- | --- | --- | --- | --- | --- | --- | --- | --- |
| Sep_2021 | 2064 | 3052 | 51.3 | 6.74 | -67 | -88 | -1.6 | -0.07 |
| Oct_2021 | 3427 | 2434 | 35.5 | 8.14 | 90 | 76 | 0.7 | 0.13 |
| Nov_2021 | 4213 | 4256 | 67.0 | 9.59 | -23 | -2 | 0.3 | -0.04 |
| Dec_2021 | 2874 | 3455 | 56.3 | 6.54 | -48 | -56 | -0.5 | -0.10 |

**Table 8.**
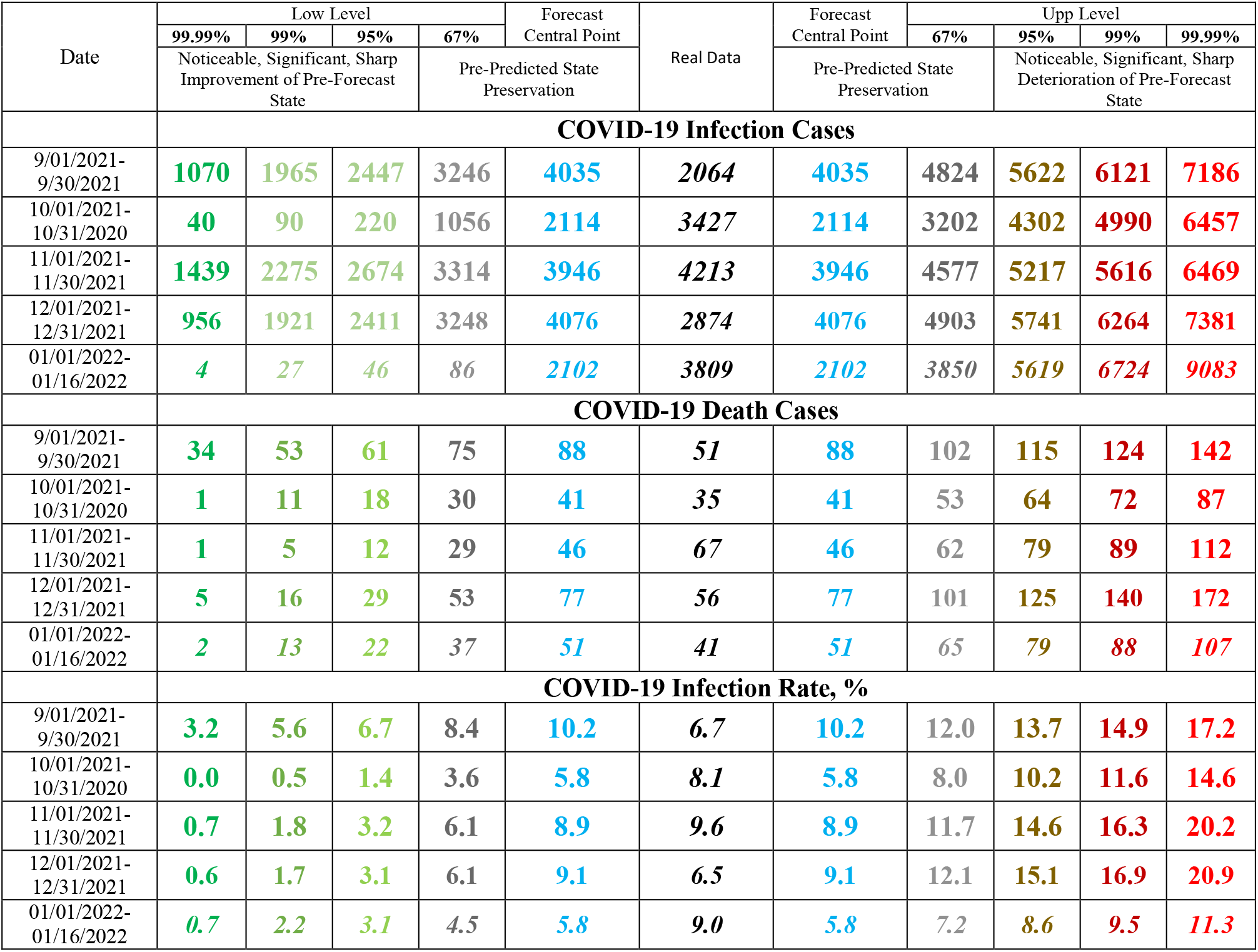
Verification of monthly interval prediction of COVID-19 confirmed infection cases, deaths cases and infection rate in Georgia from 01.09.2021 to 31.12.2021. and from 01.01.2022 to 16.01.2022.

**Table 9.** Change in the forecast state of C, D and I in relation to the pre-predicted one according to Table 1 scale [8].

| Date | COVID-19 Infection Cases |
| --- | --- |
| 9/01/2021- 9/30/2021 | Significant improvement, stability of a time-series of observations |
| 10/01/2021-10/31/2020 | Noticeable deterioration, stability of a time-series of observations |
| 11/01/2021-11/30/2021 | Preservation, stability of a time-series of observations |
| 12/01/2021-12/31/2021 | Noticeable improvement, stability of a time-series of observations |
| 01/01/2022-01/16/2022 | Preservation, stability of a time-series of observations |
| **COVID-19 Death Cases** |  |
| 9/01/2021- 9/30/2021 | Sharp improvement, stability of a time-series of observations |
| 10/01/2021-10/31/2020 | Preservation, stability of a time-series of observations |
| 11/01/2021-11/30/2021 | Noticeable deterioration, stability of a time-series of observations |
| 12/01/2021-12/31/2021 | Preservation, stability of a time-series of observations |
| 01/01/2022-01/16/2022 | Preservation, stability of a time-series of observations |
| **COVID-19 Infection Rate** |  |
| 9/01/2021- 9/30/2021 | Noticeable improvement, stability of a time-series of observations |
| 10/01/2021-10/31/2020 | Noticeable deterioration, stability of a time-series of observations |
| 11/01/2021-11/30/2021 | Preservation, stability of a time-series of observations |
| 12/01/2021-12/31/2021 | Preservation, stability of a time-series of observations |
| 01/01/2022-01/16/2022 | Significant deterioration, stability of a time-series of observations |

### 3.1 Comparison of time-series of Covid-19 confirmed and deaths cases in Georgia, its neighboring countries and World in summer 2021

The time-series curves and statistical characteristics of Covid-19 confirmed and deaths cases (to 1 million populations) in Georgia, its neighboring countries and World from September 01, 2021 to December 31, 2021 in Fig. 1 - 2 and in Table 1-2 are presented.

Variability of the values of C per 1 million populations is as follows (Fig. 1, Table 1):

- Georgia. Range of change: 178-1616, mean value - 843.
- Armenia. Range of change: 20-877, mean value - 284.
- Azerbaijan. Range of change: 0-716, mean value - 154.
- Russia. Range of change: 117-276, mean value - 197.
- Turkey. Range of change: 0-898, mean value -298.
- World. Range of change: 39-248, mean value - 73.

The largest variations in C values were observed in Azerbaijan (CV=77.1%), the smallest - in Russia (CV=24.8%).

Significant linear correlation (r_min_ = ± 0. 18, α = 0.05) between these countries on C value varies from 0.23 (pairs Georgia-Azerbaijan, Armenia-Azerbaijan, Armenia-Turkey) to 0.67 (pair Georgia-Russia). Linear correlation between World and these countries is significant only for two pairs: World-Armenia (r = -0.45) and World-Russia (r = -0.27). The degree of correlation [41] is as follows: moderate correlation (0.5 ≤ R < 0.7) – for pairs Georgia-Armenia, Georgia-Russia and Armenia-Russia; low correlation (0.3 ≤ R< 0.5) – for pair World-Armenia; negligible correlation (0 ≤ R < 0.3) – for pairs Georgia-Azerbaijan, Armenia-Azerbaijan, Armenia-Turkey and World-Russia.

Variability of the values of D per 1 million populations is as follows (Fig. 2, Table 2):

- Georgia. Range of change: 6.2-23.1, mean value – 14.1.
- Armenia. Range of change: 0.3-23.6, mean value – 8.7.
- Azerbaijan. Range of change: 0.0-10.9, mean value – 2.2.
- Russia. Range of change: 4.9-8.4, mean value – 6.9.
- Turkey. Range of change: 0.0-3.7, mean value -2.5.
- World. Range of change: 0.4-1.6, mean value – 1.0.

The largest variations in D values were observed in Azerbaijan (CV=75.3%), the smallest - in Russia (CV=16.0 %).

Significant positive linear correlation between these countries on D value varies from 0.19 (pair Azerbaijan-Turkey) to 0.56 (pair Armenia-Russia); negative – between Russia and Turkey (r = -0.33). Linear positive correlation between World and these countries is significant for two pairs: World-Azerbaijan (r = 0.20) and World-Turkey (r = 0.37); negative – for pair World-Russia (r = -0.27).

The degree of correlation [41] is as follows: moderate correlation – only for pair Armenia-Russia; low correlation – for pairs Georgia-Russia, Russia-Turkey and World-Turkey; negligible correlation – for pairs Azerbaijan-Turkey, World-Azerbaijan and World-Russia.

It should be noted that, in general, in the studied period of time, level of correlations shown in Tables 1 and 2 are worse than in the summer of 2021 [10].

In Table 3 the statistical characteristics of mean monthly values of C and D related to Covid-19 for 157 countries with population ≥ 1 million inhabitants from September to December 2021 (normed per 1 million population) is presented.

As follows from this Table range of change of mean monthly values of C for 157 countries varied from 0 (all months) to 1748 (December). Average value of C for 157 countries varied from 113 (September) to 190 (December). Value of CV changes from 145% (September) to 187% (November).

Range of change of mean monthly values of D for 157 countries varied from 0 (all months) to 21.5 (November). Average value of D for 157 countries varied from 1.6 (September and October) to 2.0 (November). Value of CV changes from 148% (September) to 198% (November).

Between mean monthly values of Deaths and Confirmed cases related to Covid-19 for 157 countries with population ≥ 1 million inhabitant linear correlation and regression are observed (Fig. 3). As follows from Fig. 3 the highest growth rate of the monthly average values of D depending on C was observed in October, the smallest - in December (the corresponding values of the linear regression coefficients).

In Table 4 data about Covid-19 mean monthly values of infection (C) and deaths (D) cases from September to December 2021 (per 1 million population) and ranking of Georgia, Armenia, Azerbaijan, Russia and Turkey by these parameters (in brackets) among 157 countries with population ≥ 1 million inhabitant are presented. The corresponding values of the deaths coefficient (DC) are also given here.

In particular, as follows from this Table, mean monthly values of C for 5 country changes from 69.7 (Armenia, December, 66 place between 157 country) to 1130.0 (Georgia, November, 6 place between 157 country).

Mean monthly values of D for 5 country changes from 1.58 (Azerbaijan, December, 39 place between 157 country) to 17.96 (Georgia, November, 2 place between 157 country).

On the whole (Table 4) in October 2021 Georgia was in the 4 place on new infection cases, and in September - in the 1 place on death. Georgia took the best place in terms of confirmed cases of diseases (thirteenth) in December, and in mortality (fifth) - in October.

Mean monthly values of DC for 5 country changes from 0.75% (Turkey, October) to 6.39 (Armenia, December).

Note that the mean values of DC (%) from September to December 2021 are: Georgia – 1.67, Armenia

– 3.05, Azerbaijan – 1.42, Russia – 3.50, Turkey – 0.83, World – 1.31 (according to data from Table 1 and 2).

In summer 2021 the mean values of DC (%) were: Georgia – 1.28, Armenia – 2.08, Azerbaijan – 0.80, Russia – 3.36, Turkey – 0.80, World – 1.79 [10]. Thus, during the study period, compared to the summer 2021, the DC values in all five indicated countries increased (in Turkey, this growth is insignificant). In the World, on the contrary, the value of DC has decreased.

### 3.2 Comparison of daily death from Covid-19 in Georgia with daily mean death in 2015- 2019 from September 1, 2021 to December 31, 2021

In Fig. 4-7 data of the daily death from Covid-19 in Georgia from October 1, 2020 to December 31, 2021 in comparison with daily mean death in 2015-2019 are presented. The daily mean death in 2015-2019 in different months are: January - 155, February – 146, March - 141, April - 137, May – 131, June – 124, July – 117, August – 120, September – 112, October - 123, November - 135, December - 144 (Fig. 4, 6).

The daily death from Covid-19 from September 1, 2021 to December 31, 2021 (Fig. 4) changes from 23 (October 11, 2021) to 86 (September 3, 2021). A comparison between the daily mortality from Covid-19 in Georgia from September 01, 2021 to December 31, 2021with the average daily mortality rate in 2015-2019 shows, that the largest share value of D from mean death in 2015-2019 (Fig. 5) was 76.8 % (September 03, 2021), the smallest 18.7 % (November 10, 2021).

The most share of mean daily mortality from Covid-19 of mean daily mortality in 2015-2019 from October 2020 to December 2021 in November was observed – 49.7% (Fig. 7).

### 3.3 The statistical analysis of the daily and decade data associated with New Coronavirus COVID-19 pandemic from September to December 2021

Results of the statistical analysis of the daily and decade data associated with New Coronavirus COVID-19 pandemic in Georgia from September 1, 2021 to December 31, 2021 in Tables 5-7 and Fig. 8 – 20 are presented.

The mean and extreme values of the studied parameters are as follows (Table 5): C - mean - 3145, range: 662 - 6024; R - mean - 3293, range: 795 - 6017; D - mean - 52, range: 23 - 86; I (%) - mean – 7.74, range: 3.56– 12.04. The range of variability for all studied parameters: 23.2% (I) ≤C**V** ≤42.5% (C).

Mean decade values of confirmed and recovered coronavirus-related cases varies within the following limits (Fig. 8): C – from 1709 (3 Decade of September 2021) to 4757 (1 Decade of November 2021); R – from 1545 (1 Decade of October 2021) to 4427 (3 Decade of November 2021).

Mean decade values of deaths coronavirus-related cases (Fig. 9) varies from 30 (1 Decade of October) to 76 (2 Decade of November).

Mean decade values of infection rate coronavirus-related cases (Fig. 10) varies from 5.38 % (3 Decade of December) to 10.55 % (1 Decade of November).

Time changeability of the daily values of C, R, D and I are satisfactorily described by the tenth order polynomial (Table 6, Fig. 11-13). For clarity, the data in Fig. 12 are presented in relative units (%) in relation to their average values.

Note that from Fig. 11 and 12, as in [8-10], clearly show the shift of the time series values of R and D in relation to C.

In Fig. 14-16 data about mean values of speed of change of confirmed, recovered, deaths coronavirus-related cases and infection rate in different decades of months from September to December 2021 are presented.

Maximum mean decade values of investigation parameters are following: V(C) = +139 cases/day (1 Decade of October), V(R) = +124 cases/day (3 Decade of October), V(D) = +1.7 cases/day (3 Decade of October), V(I) = + 0.20 %/ day (1 decades of October). Min mean decade values of investigation parameters are following: V(C) = -155 cases/day (1 Decade of September), V(R) = -114 cases/day (2 Decade of September), V(D) = -2.9 cases/day (1 Decade of September), V(I) = -0.16 %/day (2 Decade of September).

Data about mean monthly values of C, R, D, I and its speed of change in from September to December 2021 in Table 7 are presented. As follows from this Table there was an increase in average monthly values of C and I from September to November, and further decrease to December. Values of R and D decrease from September to October, increase from October to November, and further decrease to December.

The values of V(C), V(R), V(D) and V(I) changes as follows: V(C) – from -67 cases/day (September) to +90 cases/day (October); V(R) - from -88 cases/day (September) to +76 cases/day (October); V(D) – from -1.6 cases/day (September) to +0.7 cases/day (October) and V(I) - from -0.10 %/day (December) to +0.13 %/day (October).

In Fig. 17 data about connection of 14-day moving average of deaths cases due to COVID-19 in Georgia with 14-day moving average of infection rate from December 18, 2020 until December 31, 2021 are presented. As follows from Fig. 17, in general, with an increase of the infection rate is observed increase of deaths cases.

Using the data in Fig. 18, a linear regression graph between the monthly mean values of D and I is obtained (Fig. 19). As follows from Fig. 19, in general a moderate level of linear correlation between these parameters is observed. This Fig. also clearly demonstrates the anomaly high mortality from coronavirus in November 2021 with relatively low value of infection rate in comparison with December 2020, which reduces the level of correlation between D and I.

As noted in [9,10] and above (Fig. 11 and 12), there is some time-lag in the values of the time series R and D with respect to C. An estimate of the values of this time-lag for September-November and October-December 2021 is given below (Fig. 20).

Cross-correlations analysis between confirmed COVID-19 cases with recovered and deaths cases shows, that in spring 2021 the maximum effect of recovery is observed 9 and 13 days after infection, and deaths - after 12-17 days [9]. In spring 2021 the cross-correlation of the C values with the R and D values weakens completely 38-45 days after infection. Maximum values of CR were 0.81 for R, and 0.79 for D.

In summer 2021 the maximum effect of recovery is observed 19 days after infection (CR =0.95), and deaths - after 16 and 18 days (CR=0.94). In contrast to the spring of 2021, in the summer of 2021, a high level of cross-correlation of C values with R and D values to 40 days after infection is observed (CR ≥ 0.80). In summer 2021 the cross-correlation of the C values with the R and D values weakens completely 56-58 days after infection. So, in Georgia in summer 2021, the duration of the impact of the delta variant of the coronavirus on people (recovery, mortality) could be up to two months [10].

From September 1, 2021 to November 30, 2021 the maximum effect of recovery is observed on 12 and 14 days after infection (CR =0.77 and 0.78 respectively), and deaths - after 7, 9, 11, 13 and 14 days (0.70≤ CR ≤0.72); from October 1, 2021 to December 31, 2021 - the maximum effect of recovery is observed on 14 days after infection (CR =0.71), and deaths - after 9 days (RC=0.43). In Georgia from September 1, 2021 to November 30, 2021 the duration of the impact of the delta variant of the coronavirus on people (recovery, mortality) could be up to 28 and 35 days respectively; from October 1, 2021 to December 31, 2021 - up to 21 and 29 days respectively (Fig. 20).

### 3.4 Comparison of real and calculated prognostic daily and monthly data related to the New Coronavirus COVID-19 pandemic in Georgia from September 1, 2021 to December 31, 2021

With September 1, 2021, we started monthly forecasting values of C, D and I. In Fig. 21-24 and Table 8 examples of comparison of real and calculated prognostic daily and mean monthly data related to the COVID-19 coronavirus pandemic in Georgia from September 1, 2021 to December 31, 2021 are presented. Note that the results of the analysis of monthly forecasting of the values of C, D and I, information about which was regularly sent to the National Center for Disease Control & Public Health of Georgia and posted on the Facebook page https://www.facebook.com/Avtandil1948/.

Comparison of real and calculated predictions data of C, D and I shown, that monthly daily and mean monthly real values of C, D and I practically falls into the 67% - 99.99% confidence interval of these predicted values for the specified time periods (Fig. 21-24, data from https://www.facebook.com/Avtandil1948/, Table 8).

For all forecast periods (4 monthly forecasting cases and one 16-days forecasting case), the stability of real time series of observations of these parameters (period for calculating the forecast + forecast period) remained (Table 9).

Thus, the daily monthly and mean monthly forecasted values of C, D and I quite adequately describe the temporal changes in their real values.

Despite the rapid spread of the omicron COVID variant an interval forecast check of confirmed COVID-19 cases, deaths and infection rates in Georgia from 01.01.2022 to 16.01.2022 confirms the representativeness of the monthly forecast for the specified time period (Table 8 and 9) [https://www.facebook.com/Avtandil1948/]. Further monitoring will determine the representativeness of this monthly forecast for the end of January 2022 in the context of the spread of mixed strains of coronavirus (Delta and Omicron) in Georgia.

## Conclusion

In the future, it is planned to continue regular similar studies for Georgia in comparison with neighboring and other countries.

## Data Availability

All data produced are available online athttps://www.soothsawyer.com/john-hopkins-time-series-data-with-us-state-and-county-city-detail-historical/; https://data.humdata.org/dataset/total-covid-19-tests-performed-by-country;
https://stopcov.ge

https://www.facebook.com/Avtandil1948/

https://stopcov.ge

